# A multicountry trial-based and model-based economic evaluation of youth mental health interventions: a protocol for assessing the short- and long-term societal impacts of YOUTHreach interventions

**DOI:** 10.64898/2026.11.12.26367972

**Authors:** Ayshe Yaylali, Lieke Maas, David McDaid, Silvia Evers, A-La Park, Thérèse van Amelsvoort, Sophie Leijdesdorff, Matthew R. Broome, Ulrich Reininghaus, Karim Lekadir, Andreas Bechdolf, Covadonga M. Díaz-Caneja, Giovanni de Girolamo, Anna-Kaisa Oidermaa, Maria Bulgheroni, Gary Donohoe, Patrick McGorry, Mario Alvarez-Jimenez, Arne Popma, Sian Lowri Griffiths, Anita Schick, Jan R. Boehnke, Sascha Struijs, AEGEE-Europe, Ghislaine A.P.G. van Mastrigt, YOUTHreach consortium

**Affiliations:** Department of Health Services Research, Care and Public Health Research Institute (CAPHRI), Maastricht University, Maastricht, the Netherlands; Care Policy and Evaluation Centre, Department of Health Policy, London School of Economics and Political Science, London, UK; Trimbos Institute, The Netherlands Institute of Mental Health and Addiction, Utrecht, The Netherlands; Department of Psychiatry and Neuropsychology, Mental Health and Neuroscience (MHeNS) Research Institute, Maastricht University, Maastricht, the Netherlands; Institute for Mental Health, School of Psychology, University of Birmingham, Birmingham, UK; Birmingham and Solihull Mental Health NHS Foundation Trust, Birmingham, UK; Department of Public Mental Health, Central Institute of Mental Health, Medical Faculty Mannheim, Heidelberg University, Mannheim, Germany; Health Service and Population Research Department, Institute of Psychiatry, Psychology & Neuroscience, King’s College London, London, UK; ESRC Centre for Society and Mental Health, King’s College London, London, UK; German Center for Mental Health (DZPG), Partner Site Mannheim-Heidelberg-Ulm, Ulm, Germany; Artificial Intelligence in Medicine Lab (BCN-AIM), Department of Mathematics and Computer Science, University of Barcelona, Barcelona, Spain; Catalan Institute for Research and Advanced Studies (ICREA), Barcelona, Spain; Department of Psychiatry and Psychotherapy, CCM, Charité -Universitätsmedizin Berlin, corporate member of Freie Universität Berlin and Humboldt-Universität Zu Berlin, Berlin, Germany; Department of Psychiatry, Psychotherapy, and Psychosomatics Incorporating FRITZ Am Urban and Soulspace, Vivantes Hospital Am Urban and Vivantes Hospital Im Friedrichshain, Berlin, Germany; German Center for Mental Health (DZPG), Berlin-Potsdam Site, Germany; Department of Child and Adolescent Psychiatry, Institute of Psychiatry and Mental Health, Hospital General Universitario Gregorio Marañón, Instituto de Investigación Sanitaria Gregorio Marañón, CIBERSAM, ISCIII, School of Medicine, Complutense University, Madrid, Spain; IRCCS Istituto Centro San Giovanni Di Dio Fatebenefratelli, Unit of Epidemiological Psychiatry and Digital Mental Health, Brescia, Italy; Peaasi, Tallinn, Estonia; Ab.Acus SRL, Milan, Italy; School of Psychology, University of Galway, Galway, Ireland; Centre for Neuroimaging, Cognition, and Genomics (NICOG), University of Galway, Galway, Ireland; Orygen, The National Centre of Excellence in Youth Mental Health, Parkville, VIC, Australia; Centre for Youth Mental Health, The University of Melbourne, Parkville, VIC, Australia; Department of Child and Adolescent Psychiatry, University Medical Center, Amsterdam, the Netherlands; School of Health Sciences, University of Dundee, Dundee, UK; Faculty of Behavioural and Movement Sciences, Department of Clinical, Neuro and Developmental Psychology, Vrije Universiteit Amsterdam, Amsterdam, The Netherlands; AEGEE, Brussels, Belgium

**Keywords:** protocol, youth mental health, prevention, health technology assessment, real-world evidence, economic evaluation, resource use measurement

## Abstract

**Background:** Mental health problems among young people impose a burden extending beyond healthcare, including impaired health-related quality of life and high societal costs from disrupted education, lost productivity, and informal care. Youth-friendly interventions may reduce this burden by providing early support and preventing escalation. However, their broader societal economic impact remains insufficiently explored. Evaluations incorporating consequences beyond healthcare and longer time horizons can better capture their societal value.

**Methods:** This protocol outlines harmonised trial-based and model-based economic evaluations of YEAH, SELFIE, and MOST, three mental health interventions for young people aged 12–25. Each intervention will be compared separately against care as usual over 12 months, with longer-term costs and outcomes projected at 2, 3, 5, and 10 years. Multicountry participant data will be pooled, using a prespecified reference country for base-case costing and methodological guidance. A harmonised, context-specific Resource Use Measurement instrument was developed to capture healthcare and broader societal resource use consistently across participating countries and to support evaluation of the economic impact of youth mental health interventions from a multicountry perspective. Uncertainty will be assessed through sensitivity and scenario analyses.

**Discussion:** The proposed harmonised methodological framework will support consistent estimation of the short- and long-term cost-effectiveness of three youth-friendly mental health interventions. It will enable comparison of intervention-specific results generated using different trial designs and support consistent estimation of costs and outcomes across the three interventions. The context-specific Resource Use Measurement instrument will further support consistent measurement of healthcare utilisation, productivity losses, and informal care across the economic evaluations.

**Conclusion:** By integrating trial-based evidence with long-term modelling and standardised measurement of resource use, this study aims to generate robust, policy-relevant evidence to inform health technology assessment and decision-making for youth mental health interventions. The context-specific Resource Use Measurement instrument is also intended to support future economic evaluations of mental health interventions conducted across multiple countries.

## Introduction

Mental disorders commonly emerge during adolescence and early adulthood and contribute substantially to the adverse impacts of disease, with economic costs extending well beyond the healthcare sector (1–3). Despite high levels of need, young people’s access to timely and appropriate mental health care can be limited (4, 5). In many countries, young people often face uncertainty when seeking access to services. This uncertainty is compounded by limited access to support in primary care, lengthy referral processes, long waiting lists, the need to consult multiple individuals before treatment initiation, and concerns about privacy and confidentiality. Moreover, multiple barriers may prevent young people from accessing care or from remaining engaged with it (6).

Youth-friendly, low-threshold interventions delivered outside of the formal healthcare system may reduce adverse societal impacts by providing early support and reducing the risk of escalation (7). The wave of innovation in youth mental health care is spreading globally, with at least 13 countries adopting an integrated youth primary care model adapted to local cultural, health financing patterns, and workforce contexts (4). YOUTHreach aims to evaluate the clinical and cost-effectiveness of three existing innovative interventions in seven European countries: Germany, Ireland, the United Kingdom, the Netherlands, Italy, Spain and Estonia, as well as in Australia (8). These three complementary youth mental health interventions are YEAH (Youth mEntal heAltH support centres), youth-friendly Integrated Youth Mental Health Services (IYMHS) offering free and easily accessible support, generally without waiting lists or appointments (9); SELFIE (Self-Esteem Low-intensity Friendly Intervention Ecological), a transdiagnostic blended ecological momentary intervention providing personalised, real-time support to young people with low self-esteem who have experienced childhood adversity (10); and MOST (Moderated Online Social Therapy), a hybrid digital platform integrating evidence-based psychological treatment with professional and peer support within a moderated online community (11, 12). Across the three evaluations, care as usual (CAU) will comprise the health, mental health, social, and other support services ordinarily available to participants within each country and study site. Participants in the control group will continue to access these services but will not receive the intervention under evaluation. As care pathways differ across countries, the type and intensity of CAU will be documented using a context-specific Resource Use Measurement (RUM) instrument, developed for this study through a multistakeholder involvement process and incorporated into the economic evaluations (8).

SELFIE and MOST will be evaluated in a randomised controlled trial, while YEAH will be evaluated using a quasi-experimental design, reflecting its real-world, open-access implementation. The base-case economic analyses will adopt a societal perspective, while scenario analyses will examine the healthcare perspective. The societal perspective accounts for healthcare costs (e.g., hospitalisations, GP visits, medications, rehabilitation, formal caregiving, and intervention costs), costs to young people and their families (e.g., travel expenses, out-of-pocket payments, time spent receiving care, and informal caregiving time), productivity costs including absenteeism, presenteeism, and unpaid work loss, as well as costs outside the healthcare system (e.g., social care, education, and support services) (13, 14). A healthcare perspective considers only costs and outcomes occurring within the healthcare system or within the scope of the relevant healthcare budget holder, typically including healthcare resource use and direct medical costs (13).

Emerging economic evidence on youth-friendly mental health interventions is promising but remains heterogeneous. In Australia, a national evaluation of the headspace programme estimated an incremental cost-effectiveness ratio of AUD $44,722 per quality-adjusted life year (QALY) gained over 18 months, suggesting that the programme represented cost-effective care. However, results varied substantially across service locations and modelling assumptions (15). In the Netherlands, the @ease walk-in IYMHS have been shown to serve a population with a considerable illness burden, with mean health-related quality of life of 0.64 compared with 0.91–0.96 in the general Dutch youth population, and average costs of €579 per person in the three months preceding their first visit, equivalent to €2,318 per person annually (16). From a societal perspective, the Dutch SELFIE intervention had an estimated incremental cost per QALY gained of approximately €54,000. At the same time, the healthcare-perspective analysis yielded an incremental cost of €15,671 per QALY and a 77% probability of cost-effectiveness at a willingness-to-pay threshold for health care providers of € 50,000 per QALY gained (17). In Australia, an economic evaluation of Horyzons, an earlier moderated online social therapy intervention underlying MOST, found significantly lower costs than treatment as usual from both healthcare and societal perspectives and was dominant in the cost-effectiveness analysis based on social functioning, although the cost-utility analysis showed fewer QALYs and the differences in outcomes were not statistically significant (18). Together, these studies provide promising but mixed evidence on the economic value of youth mental health interventions and underline the need for harmonised evaluations across interventions and countries.

Economic evaluations conducted alongside clinical studies estimate resource use, costs, outcomes, and cost-effectiveness over an observed follow-up period using individual-level data and generally require fewer assumptions than model-based analyses (19, 20). However, trial-based economic evaluations may not capture consequences beyond the study horizon, which is often just one year or less (21). This is particularly relevant in youth mental health, as the effects of untreated or inadequately treated mental health problems may persist well beyond the study period (22). YOUTHreach will combine trial-based and model-based economic evaluations within a harmonised methodological framework. Three 12-month trial-based economic evaluations will assess YEAH, SELFIE, and MOST separately against CAU. YEAH will be evaluated using propensity score-adjusted quasi-experimental data, whereas SELFIE and MOST will be evaluated using randomised controlled trial data. These analyses will be complemented by three model-based economic evaluations projecting the costs and outcomes of the same intervention–comparator pairs over 2-, 3-, 5-, and 10-year time horizons.

For each intervention, individual-level resource-use and outcome data from all participating countries will be pooled. One participating country will be selected as the reference country for the pooled base-case valuation framework, based on prespecified criteria including participant numbers, data completeness and quality, availability of national unit costs and outcome value sets, and national health technology assessment guidance. The selected country’s costing methods, outcome valuation methods, analytical perspectives, and other relevant methodological recommendations will be applied consistently in the pooled base-case analyses. The methodological assumptions will be examined in scenario and sensitivity analyses.

Many economic evaluation methods and resource-use tools were developed mainly for adult populations and may not fully capture the broader costs and outcomes relevant to some younger people. This is particularly important in youth mental health, where impacts may extend beyond healthcare to education, social care, family support, and other sectors (23, 24). To address this gap, YOUTHreach developed a context-specific RUM instrument through a multi-stakeholder process involving young people, health economists, policymakers, psychologists and service professionals. Their input ensured the instrument reflects young people’s and service providers’ experiences and captures relevant resource use within and outside the healthcare system, including education and social support services. The development process was informed by established health-economic methods, including principles developed by the Institute for Medical Technology Assessment (iMTA) Medical Consumption Questionnaire (iMCQ) and

Productivity Cost Questionnaire (iPCQ) (25, 26), to support valid and consistent cost estimation across the YOUTHreach economic evaluations.

This study aims to establish a harmonised methodological framework for evaluating the cost-effectiveness of YEAH, SELFIE, and MOST compared with CAU among young people aged 12–25 years. Twelve-month cost-utility and cost-effectiveness analyses will be conducted using individual-level cost and outcome data, complemented by model-based analyses projecting costs and outcomes over 2-, 3-, 5-, and 10-year time horizons. The base-case analyses will be conducted from a societal perspective, with healthcare-perspective analyses conducted as scenarios. The following research questions will be addressed.

RQ1: How will the 12-month cost-effectiveness of YEAH compared with CAU be estimated using propensity score-weighted quasi-experimental data?

RQ2: How will the 12-month cost-effectiveness of SELFIE and MOST compared with CAU be estimated using randomised controlled trial data?

RQ3: How will the long-term cost-effectiveness of YEAH, SELFIE, and MOST be projected over 2-, 3-, 5-, and 10-year time horizons using the Markov model?

In addition, the study describes the multistakeholder development of a context-specific RUM instrument that captures relevant societal costs and provides translated versions for all participating countries to support its use in future studies.

## Methods

### Study design and setting

Figure 1 summarises the designs of the three multicentre, multicountry studies. YEAH is a prospective quasi-experimental cohort study with non-equivalent groups. The intervention group consists of young people presenting to a participating YEAH IYMHS for the first time. However, previous or concurrent use of counselling or mental health services is permitted. Control participants are recruited separately from the corresponding catchment areas via local mental health services, schools, student and educational counselling services, digital outreach such as targeted online advertisements and social media, and population registers. To enhance recruitment efficiency, data from 336 participants assigned to the control conditions of the SELFIE and MOST trials will also be included in the YEAH control group. Propensity score weighting will be applied to adjust for measured baseline differences between the YEAH and control groups (8, 9).

**Figure 1.**
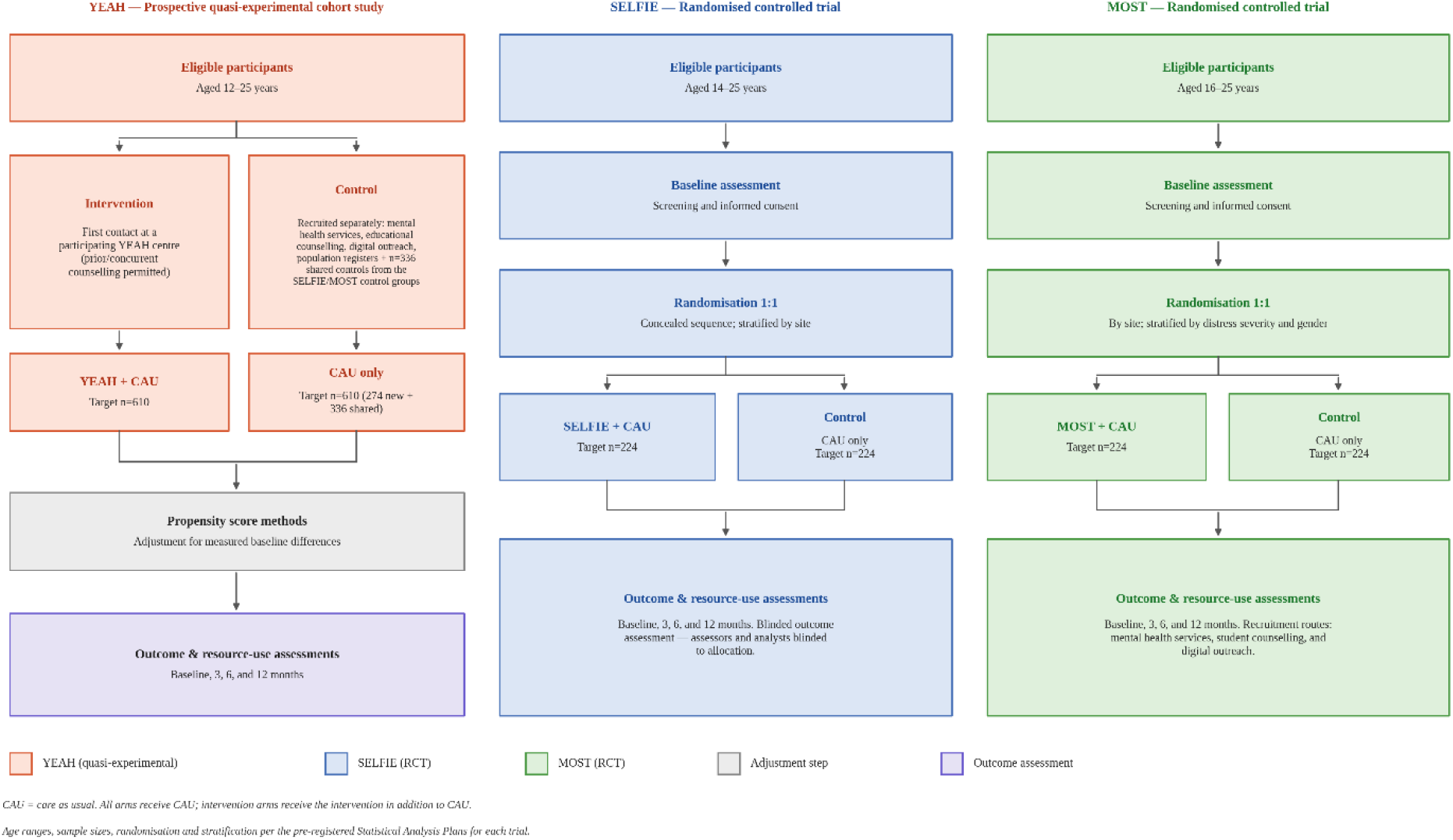
Overview of trial designs across YEAH, SELFIE, and MOST. ***Note:*** Participants in the intervention groups receive the respective YOUTHreach intervention in addition to any care as usual available to them; participants in the control groups receive care as usual alone. Therefore, the formal comparisons are YEAH plus care as usual versus care as usual alone, SELFIE plus care as usual versus care as usual alone, and MOST plus care as usual versus care as usual alone. Abbreviations: YEAH: Youth mEntal heAltH support centres; SELFIE: Self-Esteem Low-intensity Friendly Intervention Ecological; MOST: Moderated Online Social Therapy; n, target number of participants; CAU: Care as Usual

SELFIE and MOST are multicentre, multicountry, parallel-group randomised controlled trials in which eligible participants are individually randomised 1:1 to receive either the respective intervention plus CAU or CAU alone. Outcomes are assessed at baseline and at 3, 6, and 12 months of follow-up. In SELFIE, allocation is determined by a concealed randomisation sequence generated by an independent trial statistician and stratified by clinical investigation site. Outcome assessors and analysts remain blinded to allocation. In MOST, computer-generated block randomisation is conducted separately at each operational site and stratified by baseline psychological distress severity and gender. (12, 27).

CAU differs across the three interventions due to differences in target populations, recruitment settings, and service structures (8, 9, 12, 27). In YEAH, participants may receive varying levels of existing support depending on local service availability and referral pathways because controls were recruited from multiple sources, including local mental health services, school and student counselling services, digital outreach, and population registers, meaning that prior service contact ranges from no formal care at all to relatively intensive psychiatric support depending on the recruitment channel (9). In SELFIE, participants are recruited through mental health and community services and may already receive ongoing psychological or psychiatric support (27). In MOST, participants are help-seeking young people recruited through youth mental health and related services, with some receiving concurrent face-to-face care, while others are on waiting lists or approaching discharge (6, 17, 28). Across all three interventions, participants in the control group continue to receive all publicly available mental health care in accordance with local and national service guidelines throughout the study period, without access to the respective interventions (9, 12, 27). Individuals may also access mental health support services through the education system in all settings.

The countries contributing data to economic evaluation vary by intervention. Table 1 shows the countries contributing to YEAH, SELFIE and MOST and identifies where data are drawn from existing studies.

**Table 1.** Country participation in the YOUTHreach economic evaluations.

| **Study** | **Countries** | **N countries** |
|---|---|---|
| **YEAH** | Estonia, Germany, Ireland, Italy, the Netherlands, Spain, the United Kingdom and Australia* | **8** |
| **SELFIE** | Estonia, Germany, Italy, the Netherlands, Spain and the United Kingdom | **6** |
| **MOST** | Ireland, the Netherlands, Spain and Australia* | **4** |

The Australian component MOST will use data from the existing Evolve study. For the Australian YEAH analysis, the two groups come from different studies. Data for the intervention group come from the MOST Evaluation, and data for the comparison group come from the Evolve study.

### Study Status and Timeline

Recruitment for YEAH and SELFIE is expected to end in December 2027. With 12 months of follow-up, data collection for both studies is expected to be completed in December 2028. For MOST, recruitment is currently scheduled to end on 1 July 2027, with final follow-up assessments expected in July 2028. If the proposed extension to 31 December 2027 is approved, data collection is expected to be completed in December 2028.

Recruitment is ongoing, and the economic evaluations described in this protocol have not yet generated results. The trial-based economic evaluation results for YEAH, SELFIE and MOST are expected in September 2029, and the longer-term model-based economic evaluation results in December 2029, in line with the project’s planned reporting schedule.

### Participants and eligibility criteria

Eligible participants are young people aged 12–25 years seeking support for mental health problems at participating YOUTHreach sites across Europe and Australia. Eligibility age ranges differ across the three intervention studies and are defined by the respective YEAH, SELFIE, and MOST protocols: 12–25 years for YEAH, 14–25 years for SELFIE, and 16–25 years for MOST. Recruitment takes place through clinical and community-based mental health services, youth mental health support centres, student counselling services, and digital intervention platforms participating in the YOUTHreach consortium.

As the three interventions differ in design and implementation, recruitment procedures vary slightly between interventions. For the YEAH intervention, participants are recruited through IYMHS and corresponding catchment areas in participating countries which are *headspace (Australia), @ease (the Netherlands), soulspace (Germany), Jigsaw (Ireland), Peaasi (Estonia), PAUSE (United Kingdom), and Centro Ponti (Italy*). In Spain, YEAH recruitment is planned at *Hospital General Universitario Gregorio Marañón in Madrid (Servicio Madrileño de Salud)*. Young people receiving CAU within the same catchment areas are also included for comparison. For the SELFIE intervention, participants are recruited through mental health services at the participating clinical investigation sites, as well as through school psychology, student and educational counselling services, digital outreach, and population registers within the corresponding catchment areas in the Netherlands, Estonia, Germany, the United Kingdom, Spain, and Italy (8). For the MOST intervention, participants are recruited through youth mental health services, student counselling services, and targeted outreach activities, including social media recruitment, across participating sites in the Netherlands, Ireland, Spain, and Australia (8, 28).

Participants are eligible if they are seeking support for mental health problems, demonstrate clinically meaningful psychological distress according to the relevant intervention protocol, and are able and willing to provide informed consent. Psychological distress is assessed using validated screening instruments such as the CORE-10 or comparable measures, depending on the intervention protocol.

Eligibility criteria are assessed during the screening procedure before enrolment, and eligible participants complete baseline assessments before entering the study conditions (6, 8, 17). Eligible participants receive study information and provide informed consent before participating. For minors, parental or guardian consent is obtained where required, in accordance with country-specific ethical and legal requirements (8). Participants are excluded if they cannot provide informed consent, lack sufficient language proficiency to complete study assessments, or present with acute psychiatric conditions requiring immediate specialist care, making participation clinically inappropriate.

### Data collection

Participants complete electronic questionnaires at baseline and follow-up time points using secure data collection systems. Participation is voluntary, and participants may skip questions or withdraw without consequences for the support they receive. Questionnaire data are linked over time using anonymised or pseudonymised identifiers. Personal identifying information is stored separately from analytical datasets. Data are stored securely and accessed only by authorised researchers in accordance with GDPR, national regulations and institutional data management policies. Further details are described in the published YOUTHreach protocol (8).

Young people who access participating services or digital platforms receive support from trained staff or moderators, depending on the intervention. During the initial contact, staff members explore the reasons for seeking support and provide guidance or appropriate support options. Clinical professionals are available within the services or on call to provide additional support or referrals if required.

In addition to the RUM and outcomes, the first part of the questionnaire collects demographic information (e.g., age, gender, occupation, living situation, country of birth, level of education, parental mental health).

### Identification, Measurement, and Valuation of the Costs and Outcomes

A pooled economic evaluation will be conducted for each intervention using individual-level resource-use and outcome data from all participating countries. To maintain consistency in valuing costs and outcomes, one participating country will be selected as the reference country for the pooled base-case valuation framework before the economic analyses. Selection will be based on prespecified criteria, including participant numbers, completeness of follow-up, availability and quality of resource-use and outcome data, availability of validated outcome measures and national value sets, feasibility of assigning national unit costs, and the existence of national costing and health technology assessment methodological guidance. The selected reference country’s costing framework, outcome valuation methods, analytical perspectives, and other relevant methodological recommendations will be applied uniformly in the pooled base-case analyses.

Cost estimation will follow a three-step microcosting framework consisting of the identification, measurement, and valuation of costs (29). Relevant cost categories have been identified by the HTA team and through expert consultations with the Health Technology Assessment Advisory Group (HAG) and Young Advisory Group (YAG) of the YOUTHreach consortium. These categories include healthcare costs (e.g., primary care, specialist visits, diagnostic procedures, hospitalisations, medications, and rehabilitation services), costs in other sectors (e.g., educational services), non-healthcare costs (e.g., informal care), and productivity costs, including absenteeism and presenteeism related to paid work, as well as productivity losses associated with unpaid work.

The resource-use volumes, including formal and informal care utilisation, will be collected through self-report using the context-specific Resource Use Measurement (RUM) instrument, applying recall periods appropriate to each resource category. Unit costs will be assigned to the measured resource-use volumes using reference prices from the selected reference country (14, 30, 31). Medication costs will be estimated using participant-level prescription costs, where available (32). Productivity losses will be valued in accordance with the selected reference country’s pharmacoeconomic guidelines, using either the friction cost approach or the human capital approach, depending on national recommendations (29, 33).

Health outcomes will be measured using the EQ-5D-5L and CORE-10. EQ-5D-5L responses will be converted into utility values using the value set of the selected reference country, where available, and used to estimate quality-adjusted life-years. CORE-10 scores will also be analysed as a secondary clinical outcome in the cost-effectiveness analyses (34, 35). For the model-based economic evaluation, future costs and outcomes will be discounted in accordance with the national health economic guidelines of the selected reference country. Table 1 in Appendix 1 summarises country-specific HTA requirements.

### Resource use measurement

The RUM instrument was developed through a multi-step, stakeholder-informed process to comprehensively measure resource use and associated impacts among individuals aged 12–25, as shown in Figure 2. Initially, a methodological review identified established approaches for measuring healthcare and broader societal resource use. This review underscored the need for a youth-specific instrument that captures resource use and impacts relevant to this age group. Consequently, an initial draft of the RUM instrument was created.

**Figure 2.**
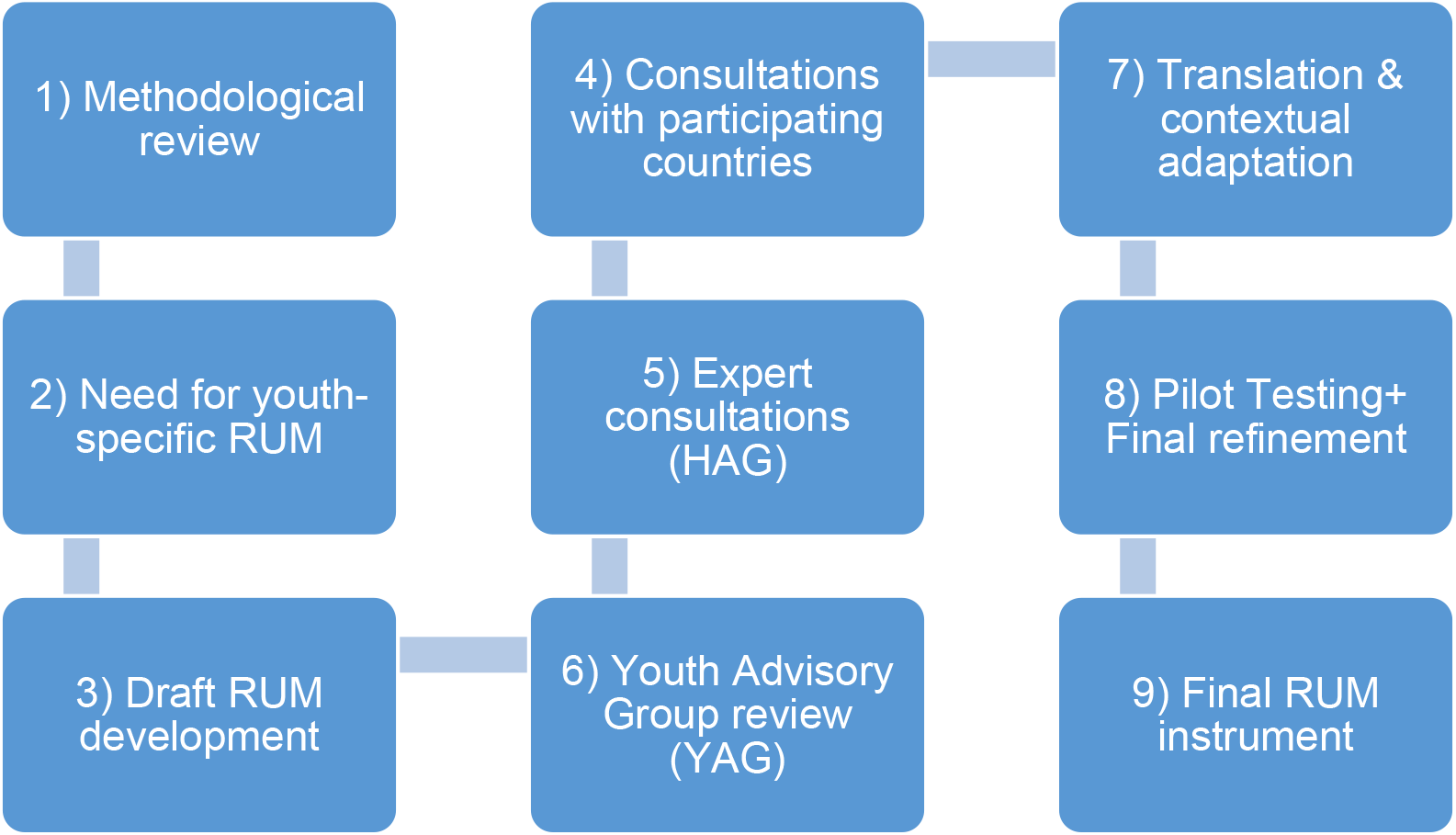
Step-by-step development of the Resource Use Measurement (RUM) instrument. HAG, Health Technology Assessment Group; YAG, Youth Advisory Group. The RUM Questionnaires for each country in their original language can be found in Appendix 2.

The draft instrument was subsequently reviewed with project partners from participating countries. These consultations identified relevant healthcare services, mental health care pathways, educational support, and other service structures within each context, while ensuring comparability across countries. Additional expert consultations were held with members of the project’s HAG, who evaluated the proposed resource-use categories, terminology, recall periods, and the instrument’s coverage of costs from both healthcare and societal perspectives. The YAG subsequently reviewed the draft instrument. Their feedback addressed the clarity, accessibility, and relevance of the questions, as well as the comprehensibility of terminology describing healthcare services, educational support, and other forms of assistance for young people.

After these consultations, the English-language questionnaire was translated into the relevant study languages, including Italian, Estonian, Spanish, German, and Dutch. The HTA team and participating country partners reviewed the translated versions to ensure linguistic accuracy, contextual appropriateness, and consistency with national healthcare and service terminology.

The instrument was subsequently pilot-tested by project partners and youth representatives. The team incorporated feedback from the translation, review, and pilot-testing stages during a final refinement phase, resulting in a context-specific RUM instrument for implementation across the YOUTHreach studies.

The instrument records contacts with healthcare professionals across primary and specialist care, including general practitioners, practice nurses, hospital-based specialists such as paediatricians, psychiatrists, psychologists, and neurologists, and community-based mental health professionals such as counsellors and psychotherapists. It also documents interactions with occupational therapists, social workers, school psychologists, and other relevant professionals.

The RUM also captures use of acute and inpatient services, including emergency department visits, ambulance transport, hospital admissions, psychiatric inpatient care, day treatment, and diagnostic visits in hospitals or mental health facilities. Additionally, it collects information on medication and supplement use, including antidepressants, attention-deficit/hyperactivity disorder medications, antipsychotics, sleep medications, and supplements.

To capture broader societal costs, the instrument includes questions about educational and workplace support services, informal care provided by friends or family, and productivity losses from school or work absenteeism and presenteeism.

### Intervention Costs

Intervention costs will be estimated using intervention-specific resource-use data collected within the YOUTHreach evaluation. The YEAH, SELFIE and MOST interventions differ substantially in delivery format, staffing requirements, and digital infrastructure, and will therefore be costed separately. Intervention costs will include relevant staff time, training and supervision, materials, and, where applicable, intervention-specific digital platform or app-related costs (8). Costs will be estimated at the individual participant level where resource-use data allow and valued using unit costs from the selected reference country.

The YEAH intervention costs reflect the IYMHS centre model. Intervention costs will include infrastructure costs such as rent, utilities, and administration, as well as staff time from the multidisciplinary teams delivering care, including psychiatrists, clinical psychologists, counsellors, social workers, peer workers, and vocational staff, estimated from existing evaluation data collected at each centre. Additional cost components include materials, training, and supervision time (6). Because the type, duration, and number of sessions vary substantially across participants and across sites, ranging from a single walk-in contact to multiple sessions of brief therapy, social counselling, job coaching, and peer support intervention, costs will be estimated at the individual participant level using the resource use measurement tool, with each service type recorded separately. Given the substantial heterogeneity in centre infrastructure and staffing models across sites, we may conduct additional sensitivity analyses alongside the reference-country base-case analysis, depending on the availability and quality of relevant information. Given this complexity, YEAH intervention cost estimates will be interpreted with caution and reported transparently, along with the assumptions made.

SELFIE intervention costs will include the platform cost associated with the TherapyDesigner system; clinician time for six intervention contacts per participant (three face-to-face sessions and three email contacts) delivered by trained mental health professionals; and costs related to the smartphone-based application. Intervention costs will include relevant one-off and participation costs, such as training and implementation, consistent with the previous SELFIE economic evaluation (17).

Intervention costs for MOST will be estimated at the individual level using a micro costing approach. The costs associated with clinician support will vary according to the assigned support intensity level, ranging from asynchronous weekly therapeutic messages and on-demand support (self-guided level) to synchronous moderator contact by phone or video of up to 20–40 minutes per week (coaching and blended teletherapy levels), consistent with previous economic evaluation of the MOST-based Horyzons intervention (18, 28). Costs will also include moderator time, peer support, vocational support (where applicable), training costs, and platform infrastructure and maintenance.

### Outcome measurement

The EQ-5D-5L, for which permission was obtained from EuroQol (36), and the Clinical Outcomes in Routine Evaluation-10 (CORE-10) are included to assess generic and mental health-specific quality of life and psychological distress, respectively.

The EQ-5D-5L is a standardised, generic instrument for measuring health-related quality of life developed by the EuroQol Group (37). It consists of five dimensions: mobility, self-care, usual activities, pain or discomfort, and anxiety or depression, each rated on five levels of severity ranging from no problems to extreme problems (38). The dedicated “anxiety/depression” dimension among its five makes the instrument a valid and widely used measure of HRQoL in mental health (39). Responses are converted into utility scores using country-specific value sets, where scores range from less than zero, representing health states considered worse than death, to one, representing full health (40). In the pooled base-case analysis, EQ-5D-5L health states will be converted into utility values using the value set of the selected reference country. Where sufficient data, time, and methodological guidance are available, country-specific EQ-5D-5L value sets may also be applied to participants from the respective countries. The approach to utility valuation will be finalised and prespecified before the economic analyses.

The CORE-10 is a brief 10-item self-report measure assessing psychological distress over the previous week. Items evaluate symptoms and functioning, including anxiety, depression, trauma, relationships, and risk, and are rated from 0 (“not at all”) to 4 (“most or all of the time”). Total scores range from 0 to 40, with higher scores indicating greater psychological distress; a score of 11 or above indicates clinically significant distress. The CORE-10 has good internal consistency (α=0.90), strong convergent validity with the CORE-OM, and demonstrated psychometric performance among older adolescents and young adults attending community mental health services (35, 41). Due to its brevity and applicability across settings, it was selected as the harmonised secondary clinical effectiveness outcome for the cost-effectiveness analyses of YEAH, SELFIE, and MOST, and is collected at baseline and at 3-, 6-, and 12-month follow-up.

### Trial-based economic evaluation

The trial-based economic evaluation will use individual-level data on costs and outcomes (20) collected over a 12-month study period. The primary outcome of the economic evaluation will be a cost-utility analysis, in which outcomes will be expressed as quality-adjusted life-years derived from the EQ-5D-5L. There will also be a secondary cost-effectiveness analysis, in which outcomes will be expressed as changes in psychological distress measured using the CORE-10. The comparator is CAU. The base-case analyses will adopt a societal perspective, while scenario analyses will examine the healthcare perspective.

These analyses will cover a 12-month follow-up period (t3) and will incorporate outcome data collected at baseline, 3 months (t1), 6 months (t2), and 12 months (t3). As shown in Figure 3, resource use is not directly measured from months 6 to 9. An appropriate within-trial adjustment will therefore be applied to estimate costs over the complete 12-month follow-up period.

**Figure 3.**
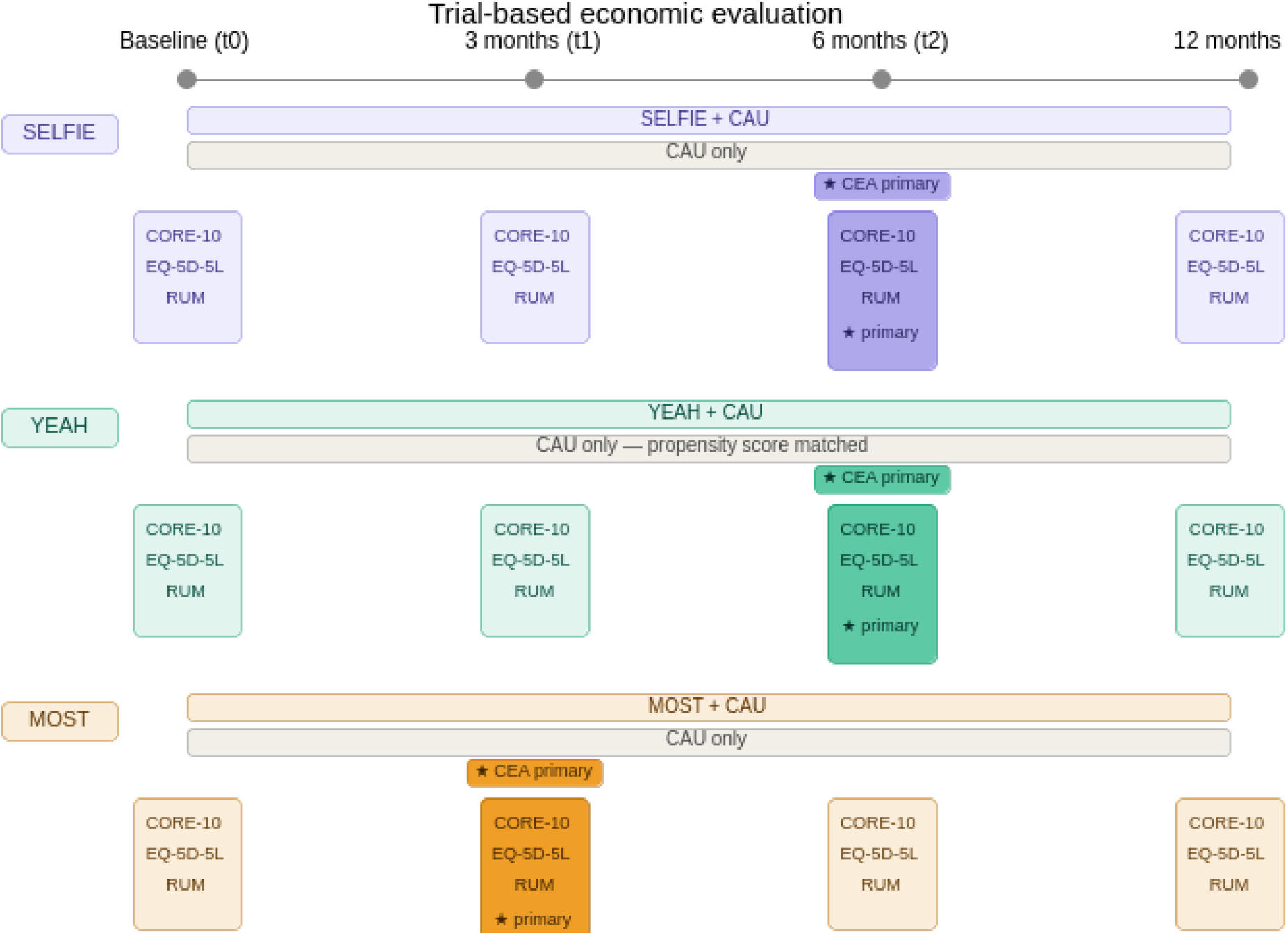
Economic evaluation assessments and primary analysis period across YEAH, SELFIE, and MOST. Notes: Resource use measurement (RUM), CORE-10, and EQ-5D-5L collected at baseline (t0), 3 months (t1), 6 months (t2), and 12 months (t3) for intervention and control groups across SELFIE, YEAH, and MOST. ★ denotes the primary cost-effectiveness analysis time period.

For YEAH, given the quasi-experimental design, propensity score weighting will be applied within the multilevel framework to adjust for systematic baseline differences between the intervention and control groups arising from non-random group formation. Results from YEAH will be interpreted as exploratory, given the inherent limitations of non-randomised designs for causal inference (42–44). SELFIE and MOST are multicentre randomised controlled trials, with individual-level randomisation within each site. To account for the resulting hierarchical/multilevel data structure, the economic evaluation will use multilevel mixed-effects regression models (45).

### Model-based economic evaluation and return on investment tool

In addition to the trial-based analyses, a model-based economic evaluation will be conducted to estimate the long-term costs and benefits of scaling up access to the interventions if shown to be effective. Again, the comparator for each intervention in the model is CAU. The model-based economic evaluation will use a probabilistic Markov decision analytic model to extrapolate impacts on resource use and outcomes beyond the observed 12-month trial follow-up period. The model will be populated using individual-level data from the trial-based economic evaluation, published literature, and expert input from the HAG and YAG through several online meetings. Trial evidence may not highlight key factors that influence implementation in different contexts. One way to capture this information is through a theory of change process, which explains how an intervention is expected to achieve its impact (46, 47). A theory of change approach in these meetings will be used to identify and document factors around engagement and pathways of care, key outcomes and impacts, and barriers and facilitators to successful implementation. This will be an iterative process with opportunities for feedback and revision as the theory of change analysis develops. The effectiveness evaluation will inform the final model time horizons; costs and outcomes are anticipated to be projected over 2, 3-, 5-, and 10-year time horizons.

Transition probabilities between health states will be estimated from observed individual-level state transitions from baseline to the three-, six-, and twelve-month follow-up time points in the trial, separately for the intervention and control arms of each trial. Health state utility values will be derived by mapping EQ-5D-5L scores to CORE-10 severity categories using observed data from trial participants. Intervention costs will be carried forward from the trial-based microcosting as described in the intervention costs section. The persistence of treatment effects beyond the trial follow-up period represents a key structural assumption. In the base case, treatment effects will be assumed to wane linearly over two years after the end of the intervention, then return to natural-history transition probabilities thereafter. The persistence assumption will be varied in scenario analyses, including full persistence of effects and immediate return to natural history at trial end.

Internal validation will be conducted by comparing model-predicted outcomes over the 12-month trial period with observed outcomes to verify that the model accurately reproduces the trial data. External validity will be assessed by comparing model predictions with published epidemiological data on the natural history of psychological distress in young people. We will consult with the HAG to obtain their feedback on model structure and preliminary results. Model development will be informed by an iterative consultation and feedback process with relevant clinical, methodological and stakeholder experts, while model validation and reporting will follow ISPOR-SMDM good research practices for modelling studies (48).

A return on investment (ROI) modelling tool that compares the cost of investment in the three interventions with the monetary value of costs averted, and any improvement in quality of life, will also be created. Policymakers and other stakeholders can use this tool to identify the long-term return on investment of the interventions (49). End users will be able to vary some of the model assumptions; for instance, this may include changes to the rates of uptake and continued engagement with interventions, implementation costs, discount rates, and the persistence of effects on quality of life and mental health. This also means that parameters can be varied in the ROI analysis to account for country-specific factors, such as differences in cost-per-QALY thresholds.

### Statistical Analysis

In line with the effectiveness studies, the economic analyses will follow the intention-to-treat (ITT) principle, including all participants in the groups to which they were originally assigned. Missing data will be handled using multiple imputation under the missing-at-random (MAR) assumption (50, 51). The final imputation approach, including the variables in the imputation model, the number of imputations, and any eligibility requirements for inclusion in the analysis, will be prespecified in the statistical analysis plan, informed by the observed data structure and relevant methodological guidance. Any deviation from the full ITT population will be reported and justified transparently. Estimates derived from multiply imputed datasets will be combined using Rubin’s rules (52, 53). For YEAH, given the higher expected attrition rate of approximately 30 per cent at six months, the imputation model will additionally include the recruitment channel as an auxiliary variable to account for systematic differences in dropout across control-group sources.

The model-based and trial-based economic evaluations will present the expected incremental cost per QALY and per clinical outcome (CORE-10) of each intervention compared to CAU. QALYs will be calculated using the area-under-the-curve (AUC) approach based on EQ-5D-5L utility values measured at baseline and at 3, 6 and 12 months. Baseline utility differences will be accounted for where appropriate (54).

For the trial-based economic evaluations, uncertainty in the incremental cost-effectiveness ratios will be characterised using non-parametric bootstrapping with 5,000 replications. The resampling approach will reflect the design and relevant hierarchical structure of each intervention study. Results will be presented on the cost-effectiveness plane and summarised using cost-effectiveness acceptability curves. For model-based economic evaluation, to explore the robustness of the results, deterministic and probabilistic sensitivity analyses will be performed (55). The model will be developed in a flexible structure that allows the incorporation of country-specific parameters where appropriate. The country-specific parameter differences are highlighted in Appendix Table 1.

### Sensitivity, Subgroup and Scenario Analysis

A series of sensitivity analyses will be conducted to assess the robustness of the base-case incremental cost-effectiveness results (19, 56). The primary analysis will use unit costs from the selected reference country, identified using the prespecified selection criteria described earlier. First, all unit costs will be varied by 10% and 20% above and below their base-case values to assess the impact of uncertainty in unit cost estimates on the incremental cost-effectiveness ratio. Where sufficient information is available, additional sensitivity analyses will examine the influence of alternative assumptions regarding intervention-related resource use and cost valuation across YEAH, SELFIE, and MOST. Second, a complete-case analysis will be conducted, including only participants with observed cost and outcome data at all four time points (53), alongside the primary analysis using imputed data, which uses all available data under the MAR assumption (51). Third, if the reference country value sets are used as the primary analysis, they will be re-estimated using an alternative EQ-5D-5L value set to assess sensitivity to the choice of utility tariff. Fourth, results will be re-estimated from a healthcare payer perspective, excluding productivity losses, informal care costs, and patient and family out-of-pocket costs, to assess how results change depending on whose costs are counted (57, 58). For YEAH specifically, a sensitivity analysis will exclude the n=336 borrowed controls from SELFIE and MOST to assess whether the base-case incremental cost-effectiveness estimate is driven by including participants recruited under different eligibility criteria.

Subgroup cost-effectiveness analyses will be conducted for predefined vulnerable groups identified in the YEAH and MOST protocols and applied consistently across all three trials including children of parents with a history of mental health problems, migrants and refugees, care-experienced young people, including care leavers, young people with disabilities, and young people identifying as LGBTQ+. These subgroups reflect the YOUTHreach consortium’s equity focus (8). Subgroup analyses will use the same multilevel mixed-model framework as the primary analysis, with an additional interaction term between condition and subgroup membership. Given the exploratory nature of subgroup analyses and the reduced sample sizes within each subgroup, results will be interpreted with caution and presented with appropriate uncertainty intervals. These analyses are pre-specified but hypothesis-generating rather than confirmatory.

Scenario analyses for the model-based economic evaluation are planned across four broad dimensions, in line with ISPOR-SMDM modelling good research practices (59). They may be refined as the model is developed. These are anticipated to include: alternative time horizons (e.g., 2, 3, 5, and 10 years) alongside the base-case, to examine how the incremental cost-effectiveness estimate evolves over different planning horizons; re-estimation from a healthcare payer perspective, excluding productivity losses, informal care costs, and patient and family out-of-pocket costs (57, 60); alternative costing assumptions, discount rates applied to future costs and effects; and alternative structural assumptions regarding the persistence of treatment effects, including full persistence beyond the trial period and immediate return to natural history transition probabilities at trial end, in place of the base-case linear waning assumption.

### Ethics, data protection, and governance

Participants provide written or electronic informed consent before participation after receiving detailed information about the study procedures and research objectives. Contact details (email) are collected only when participants opt into re-contact for future research. Personal identifying information required for study administration is stored separately from research data. Before analysis, datasets are pseudonymised to protect participant confidentiality.

Data will be processed in accordance with the General Data Protection Regulation, applicable national legislation, and the data-management policies of the participating institutions. Access to identifiable information is restricted to authorised study personnel only. Participants may withdraw until data analysis begins, as outlined in the patient information form.

## Discussion

This protocol describes a harmonised approach for conducting trial-based and model-based economic evaluations across a multicountry project. By integrating evidence generated across sites, the study aims to produce policy-relevant cost-effectiveness estimates for three youth mental health interventions that are both methodologically consistent and informative for assessing the potential transferability of findings across different healthcare systems. As part of this work, we also provide a context-specific RUM tool, applicable beyond this study, that captures costs extending beyond the healthcare sector.

### Strengths

A key strength of this protocol is its combination of trial-based and model-based approaches to capture both short-and long-term cost-effectiveness of YEAH, SELFIE, and MOST. The trial-based evaluation offers high internal validity by estimating costs and outcomes directly from observed individual-level data over the 12-month follow-up, minimising assumptions about resource use and health trajectories (20, 21). The complementary Markov model projects costs and outcomes over 2, 3, 5, and 10 years, generating long-term estimates directly relevant to HTA decision-making in each participating country. Short-term generalisability is necessarily limited by sample size and the context-specific nature of the three interventions, which are not directly comparable; the long-term projections address this by enabling deterministic and probabilistic sensitivity analyses, offering value to decision-makers seeking both local and comparative insight.

The context-specific RUM instrument is a further methodological strength. Existing economic evaluations of youth mental health interventions have been hampered by the lack of standardised resource-use tools adapted to young people’s service-use patterns and cost categories (23). The YOUTHreach RUM was developed through a rigorous, multi-step, stakeholder-informed process involving health economists, clinical experts, country partners, the HTA Group, and the Youth Advisory Group, ensuring it captures both healthcare and broader societal costs relevant to 12–25-year-olds across diverse European and Australian healthcare systems. Including education-related contacts, workplace support services, and informal care alongside standard healthcare resource use represents an advance over previous instruments in this population.

The use of EQ-5D-5L and CORE-10 as harmonised utility and effectiveness denominators across all three trials is a deliberate, methodologically justified choice, enabling consistent comparisons of cost-utility and cost-effectiveness. Where appropriate, multilevel mixed-effects regression models will be used to account for the hierarchical structure of participants nested within sites and countries. The final model specification will reflect the design and data structure of each intervention study (45, 61).

The multicountry design itself presents both an opportunity and a challenge: it strengthens external validity by capturing heterogeneity in clinical practice, unit costs, and health system structures, but introduces methodological complexity from differences in data availability, costing methods, outcome valuation, and willingness-to-pay thresholds across countries. This protocol addresses these challenges through a standardised data collection instrument, predefined analytical guidelines, and country-specific adaptations where necessary. Nonetheless, residual heterogeneity may persist and should be interpreted carefully when comparing base-case and country-specific sensitivity analysis results.

### Limitations

Integrating decision-analytic modelling within the model-based economic evaluation also requires several assumptions, both structural and parametric. These include assumptions about long-term effectiveness and cost trajectories. Although sensitivity analyses will address this uncertainty, the validity of the model-based projections ultimately depends on the quality and completeness of available data. In multicountry settings, variations in epidemiological and cost data may further amplify uncertainty. Transparent reporting and validation of the model structures are therefore essential to support credibility and reproducibility (60). Several methodological limitations should be acknowledged. First, the quasi-experimental design of YEAH represents the most significant analytical challenge in this evaluation. Because young people presenting to IYMHS cannot ethically be randomised to a waitlist or no-treatment control, the YEAH control group is recruited separately, through multiple channels, from the same catchment areas, resulting in systematic differences in help-seeking behaviour, prior service use, and baseline severity that propensity score weighting cannot fully address (42–44).

The reliance on self-reported resource use data introduces potential recall bias, particularly given the three-month retrospective recall period used in the RUM. This is a known limitation of trial-based economic evaluation and is common to all studies using self-report resource use measurement (29). The use of a stakeholder-developed, youth-friendly instrument with clear and accessible language for each service type should mitigate but not eliminate this limitation.

A further limitation of multicountry economic evaluation is the absence of fully harmonised methodological guidance across national healthcare systems. Participating countries differ in their recommended analytical perspectives, discount rates, outcome valuation methods, willingness-to-pay thresholds, and unit-cost sources, as summarised in Appendix 1. These differences limit the extent to which one set of pooled cost-effectiveness estimates can directly represent every participating country. To maintain internal consistency, individual-level data from all participating countries will be included in pooled analyses, while one country will be selected as the reference country for the pooled base-case costing, outcome valuation, and methodological framework. Uncertainty in unit costs will be examined by varying them by 10% and 20% above and below their base-case values, while alternative EQ-5D-5L outcome value sets, discount rates, and other methodological assumptions will be explored where relevant. These analyses will assess the robustness and potential transferability of the pooled findings but should not be interpreted as complete country-specific economic evaluations.

### Comparison with Existing Evidence

Previous economic evaluations of youth mental health interventions have predominantly been conducted in single countries, most often Australia, with limited transferability to European healthcare systems (17, 18, 28). The headspace model in Australia that is one of the participating IYMHS of YEAH has been the subject of the most extensive evaluation to date, demonstrating high satisfaction rates and improvements in access and functioning, but economic evidence remains limited particularly with respect to long-term cost-effectiveness and societal costs (28). In Europe, evaluations of similar services such as Jigsaw in Ireland, @ease in the Netherlands and soulspace in Germany have provided important descriptive and pre-post data, but none have incorporated formal economic evaluation with a control group or modelled long-term outcomes (6). The YOUTHreach economic evaluation addresses these gaps by providing a controlled, multicountry, multiperspective economic evaluation framework for three distinct youth mental health service models simultaneously.

The previous SELFIE economic evaluation, conducted alongside the Dutch multicentre RCT, provided initial evidence on the cost-effectiveness of SELFIE from both societal and healthcare perspectives (17). The present multicountry economic evaluation extends this evidence to participating countries with greater population diversity and heterogeneity of healthcare systems, providing a broader basis for assessing the potential transferability and policy relevance of the findings across contexts.

### Policy Relevance

Mental health problems in young people represent one of the leading causes of disease in high-income countries, and early intervention has been identified as a priority investment for reducing long-term societal costs (2, 4, 7). Decisionmakers require economic evidence that is relevant to their national health and social care contexts, but such evidence remains limited for youth mental health interventions. This evaluation will provide pooled base-case cost-effectiveness estimates for YEAH, SELFIE, and MOST using a consistent reference-country valuation framework. Sensitivity and scenario analyses will examine the influence of uncertainty in unit costs, outcome value sets, discount rates, and other key assumptions. The accompanying return-on-investment tool will allow policymakers to vary selected parameters, including intervention uptake, implementation costs, discount rates, and persistence of effects, to explore the potential relevance of the findings to their own settings.

The inclusion of vulnerable subgroups including children of parents with mental health problems, migrants and refugees, care-experienced young people, young people with disabilities and those identifying as LGBTQ+ reflects the equity focus of the YOUTHreach consortium and ensures that cost-effectiveness evidence is available for the populations most at risk of mental health problems (2) and barriers to accessing care (8).

## Conclusion

This protocol provides a robust and transparent framework for conducting economic evaluations of YEAH, SELFIE, and MOST in a multicounty setting, combining 12-month trial-based analyses with model-based projections over 2, 3-, 5-, and 10-year time horizons. The base-case analyses will adopt a societal perspective, while the healthcare perspective will be examined through scenario analyses. The accompanying resource use measurement (RUM) tool, developed specifically for this population, is a key part of this framework, enabling standardised, comparable cost data collection across countries and providing a reusable instrument for future youth mental health economic evaluations. While methodological challenges are inevitable, the proposed strategies aim to mitigate these limitations and enhance the usefulness of the results not only for decision-makers, but also for other stakeholders, including young people, researchers, and healthcare workers. Future work will focus on implementing this protocol and disseminating findings, including the RUM tool, to maximise their impact on policy and youth mental health. The broader societal economic impacts of youth mental health interventions remain insufficiently understood. By extending the analytical perspective beyond healthcare and incorporating education, employment, informal care, and longer-term outcomes, the proposed evaluations will provide a more comprehensive assessment of these interventions.

## Data Availability

No datasets were generated or analysed during the current study. All relevant data from the YOUTHreach economic evaluations will be made available upon study completion, in accordance with applicable ethical, legal, and data-protection requirements.

## Acknowledgements

We would like to acknowledge all the staff of the participating centres of YEAH: headspace (Australia), @ease (the Netherlands), soulspace (Germany), Jigsaw (Ireland), Peaasi (Estonia), PAUSE (United Kingdom), and Centro Ponti (Italy). We would like to thank the members of the YOUTHreach Health Technology Assessment Advisory Group (HAG) and Youth Advisory Group (YAG) for their valuable contributions to the development and refinement of the context-specific Resource Use Measurement instrument and for their input on the relevance, clarity, and applicability of the economic evaluation framework. We specifically acknowledge the contributions of the YAG members Alessandra Soto Ruiz and Gieli Jan van der Laan, and the HAG members Hareth Al-Janabi, Ann John, Brendan Kennelly, Adrienne Alayli, Lorena Dini, Talitha Feenstra, Ben Wijnen, Federico Tedeschi, Cathy Mihalopoulos, and Carlos García Alonso.

## Appendices

## Appendix 1: Country-specific Health Technology Assessment Requirements

**Table A1.**
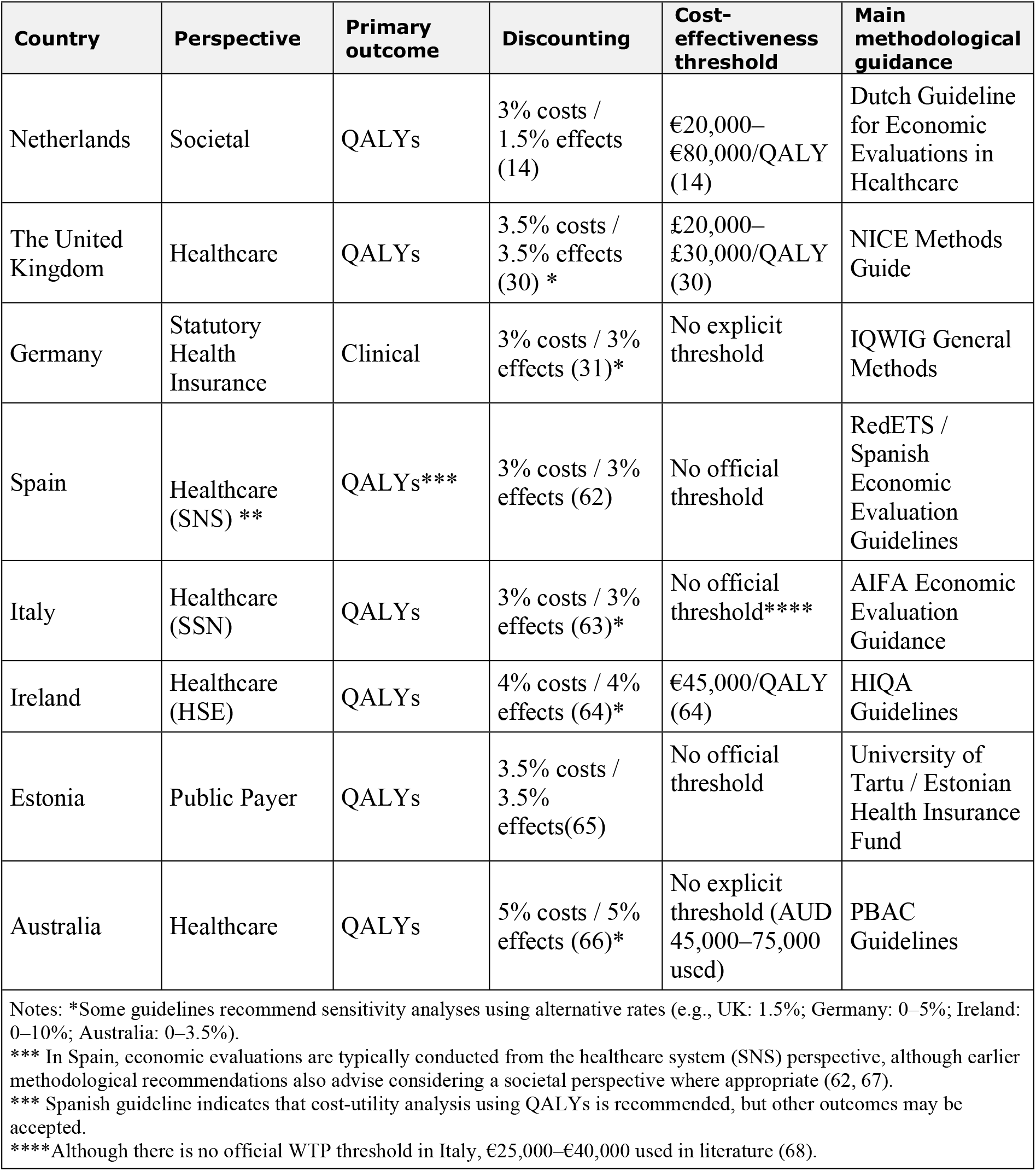
Comparison of health technology assessment requirements across participating countries.

## Appendix 2: Context-Specific Resource Use Measurement Survey for each Country included in the Study

### 2.1 Resource Use Measurement Survey for England

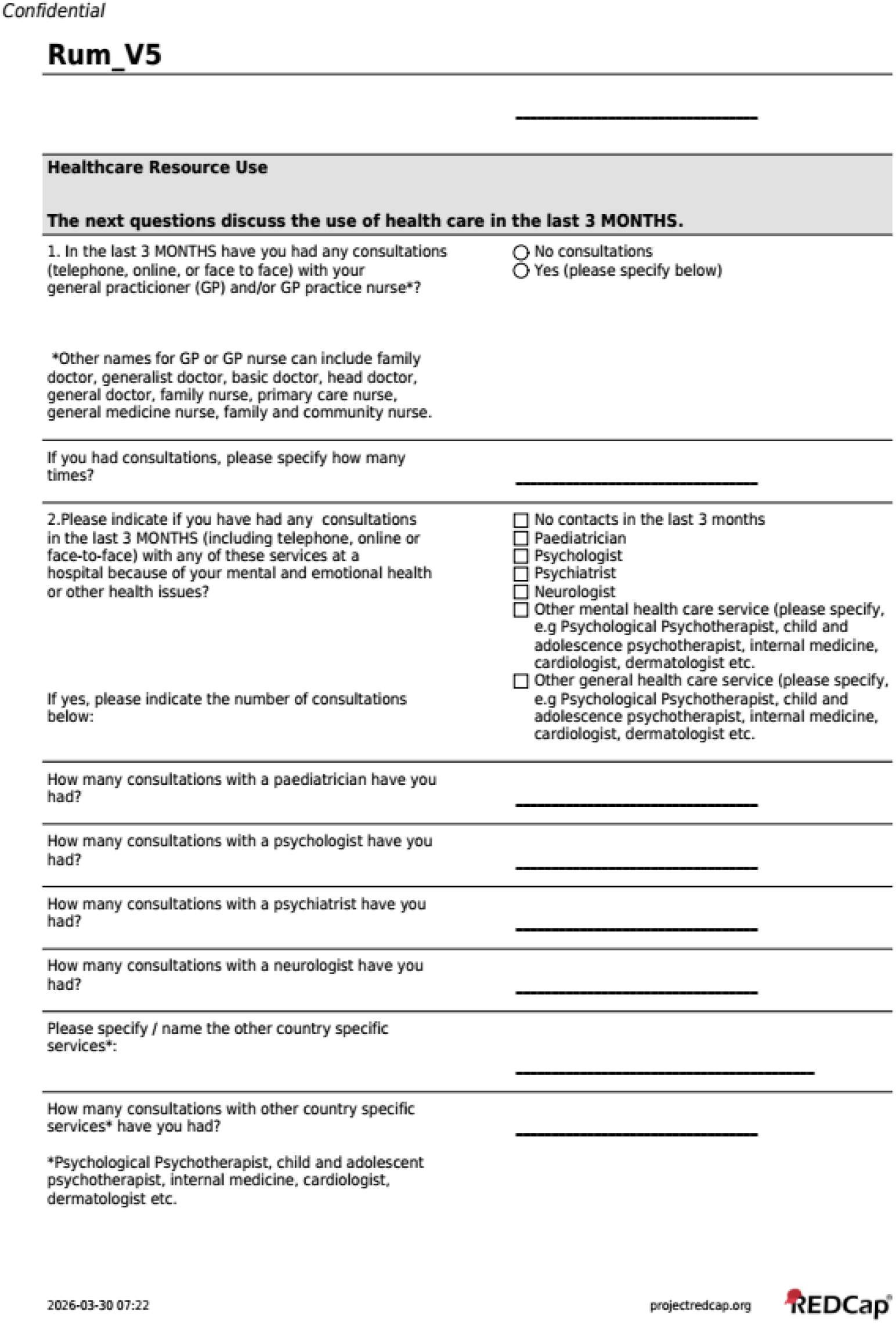
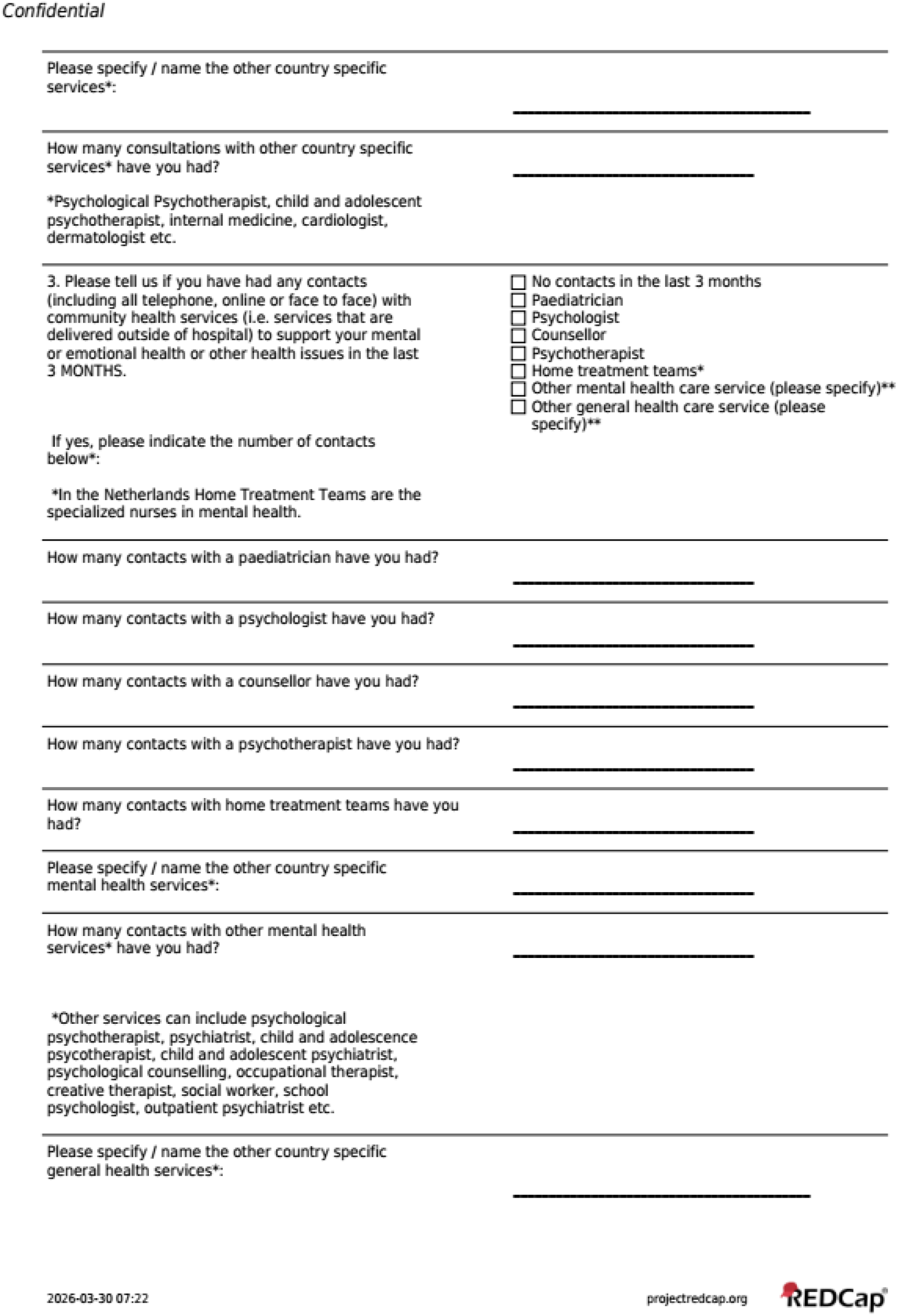
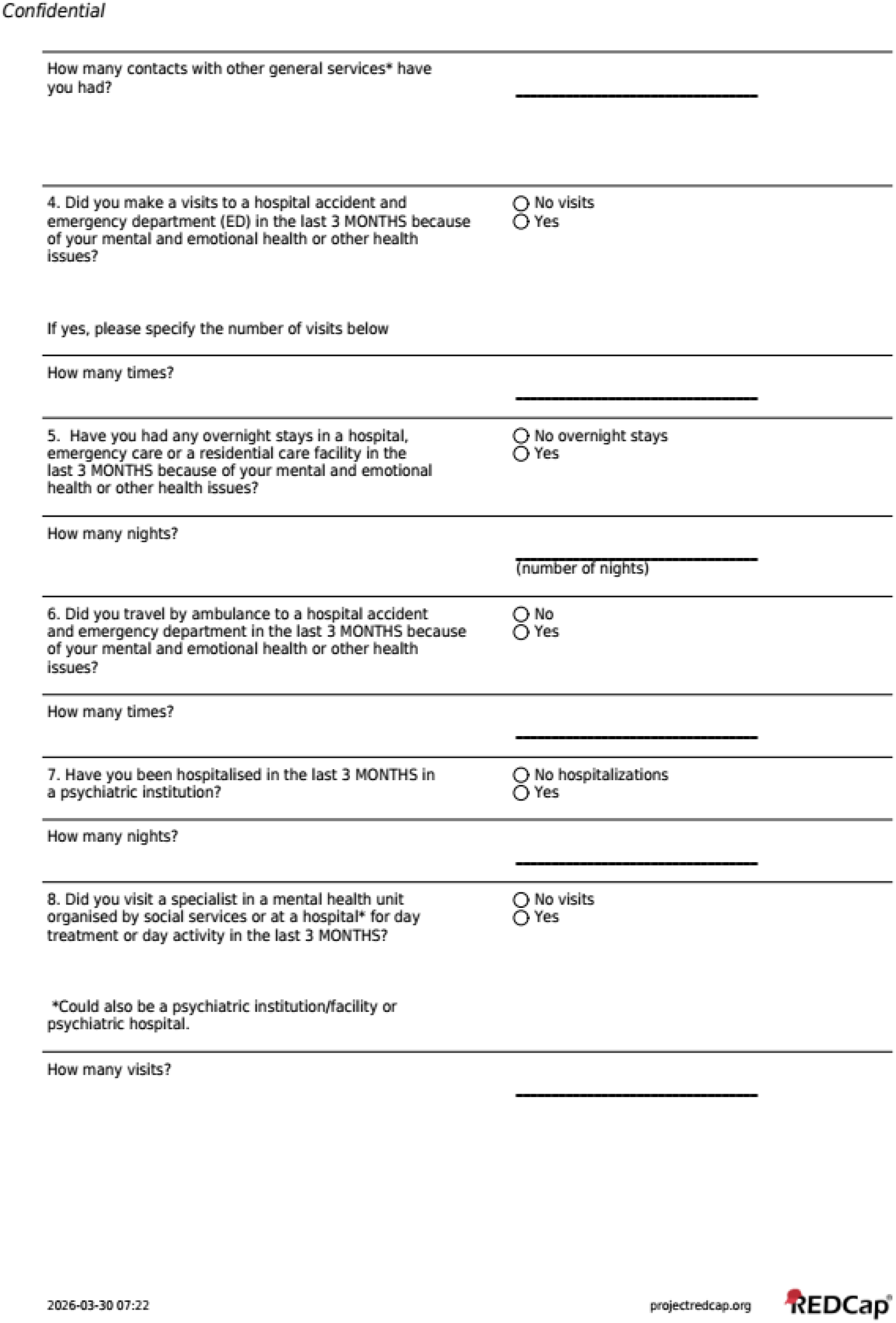
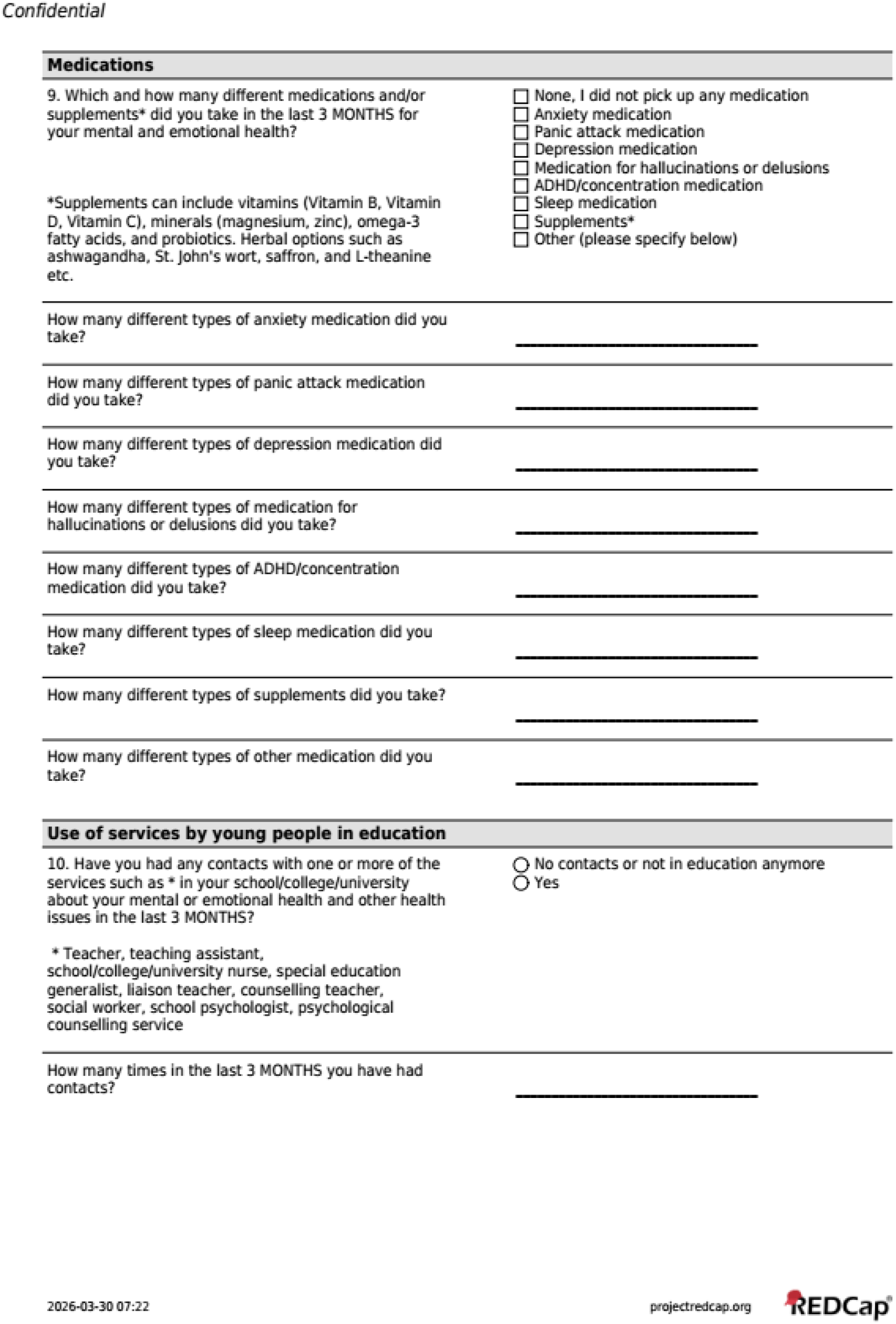
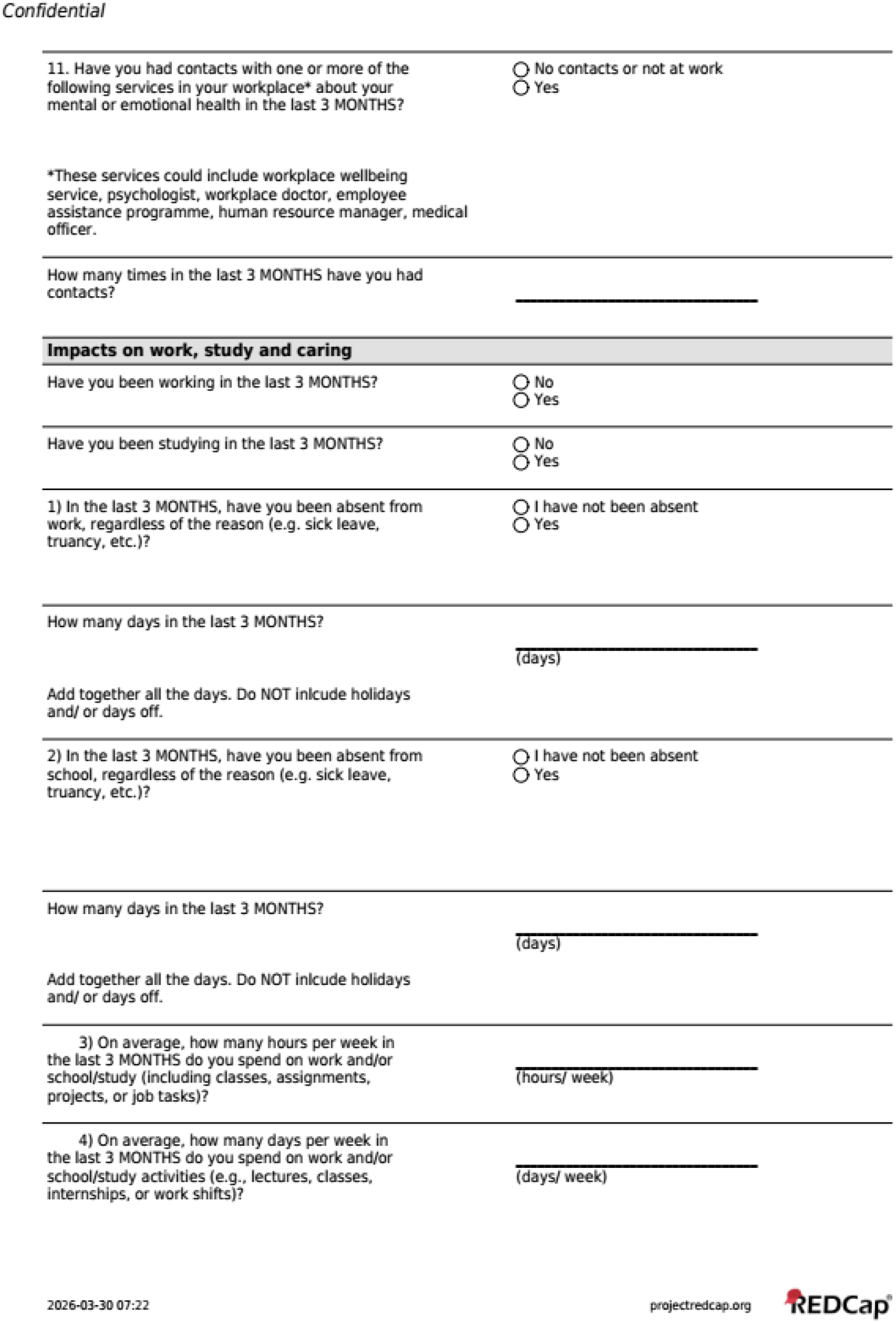
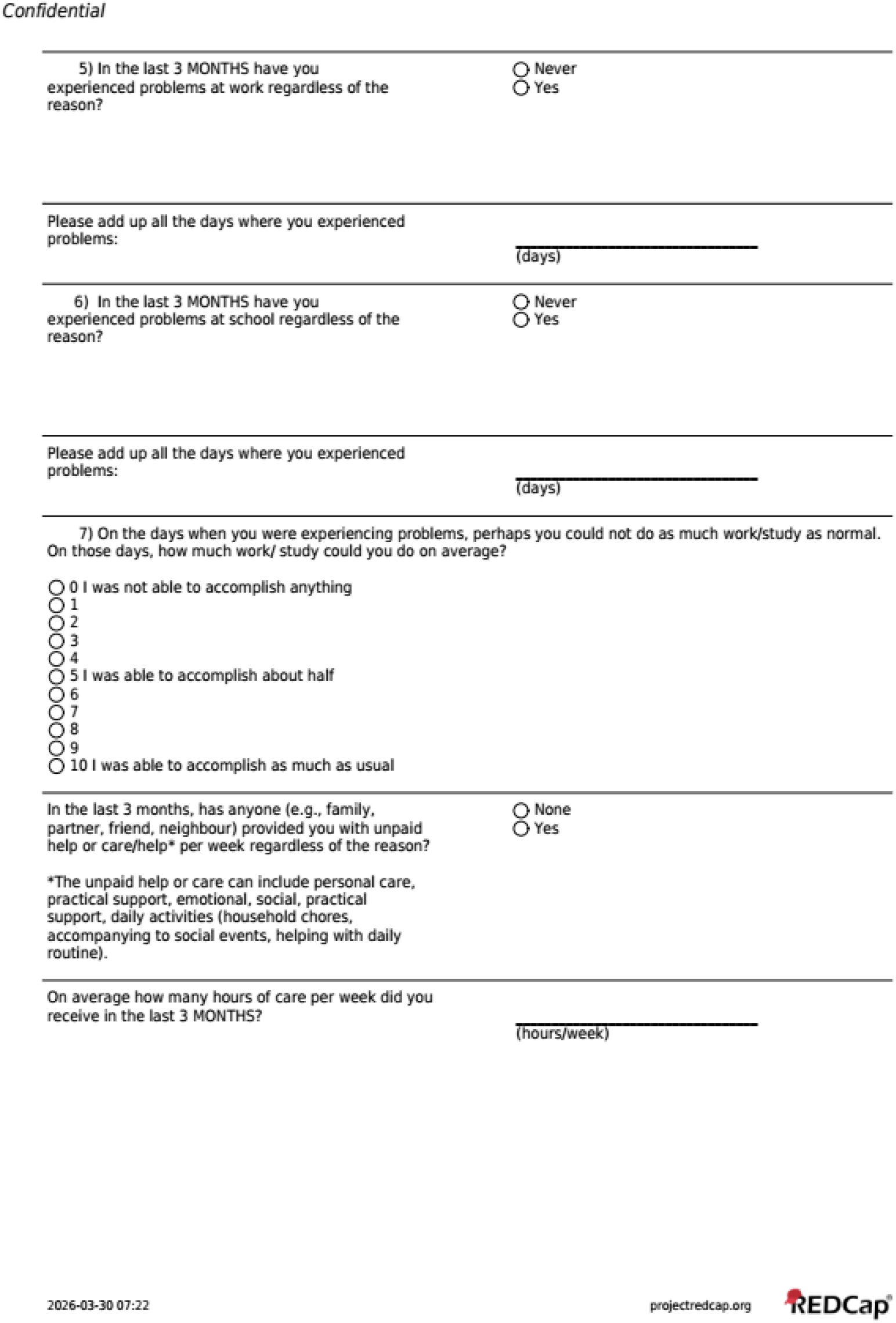

### 2.2 Resource Use Measurement Survey for Ireland

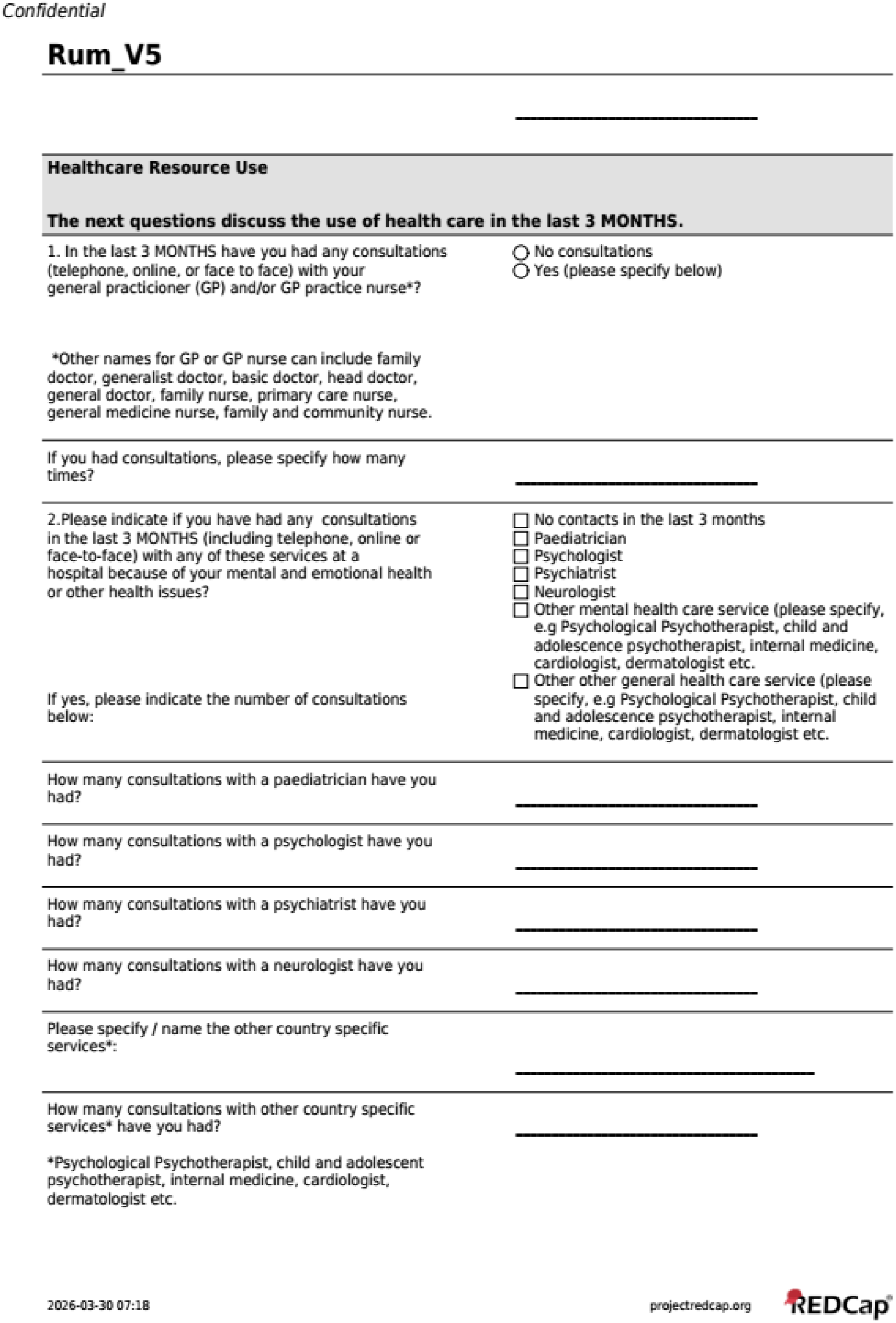
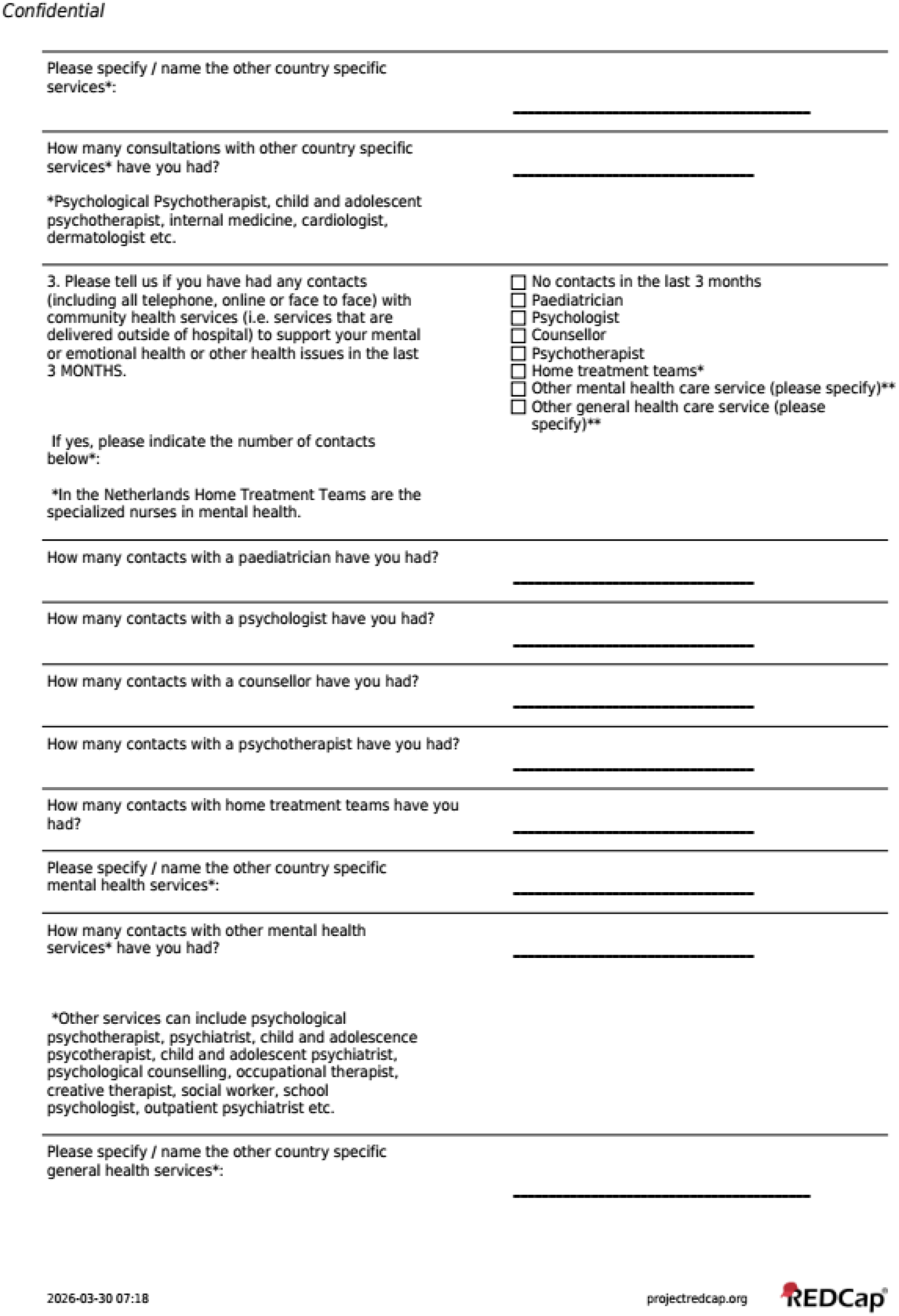
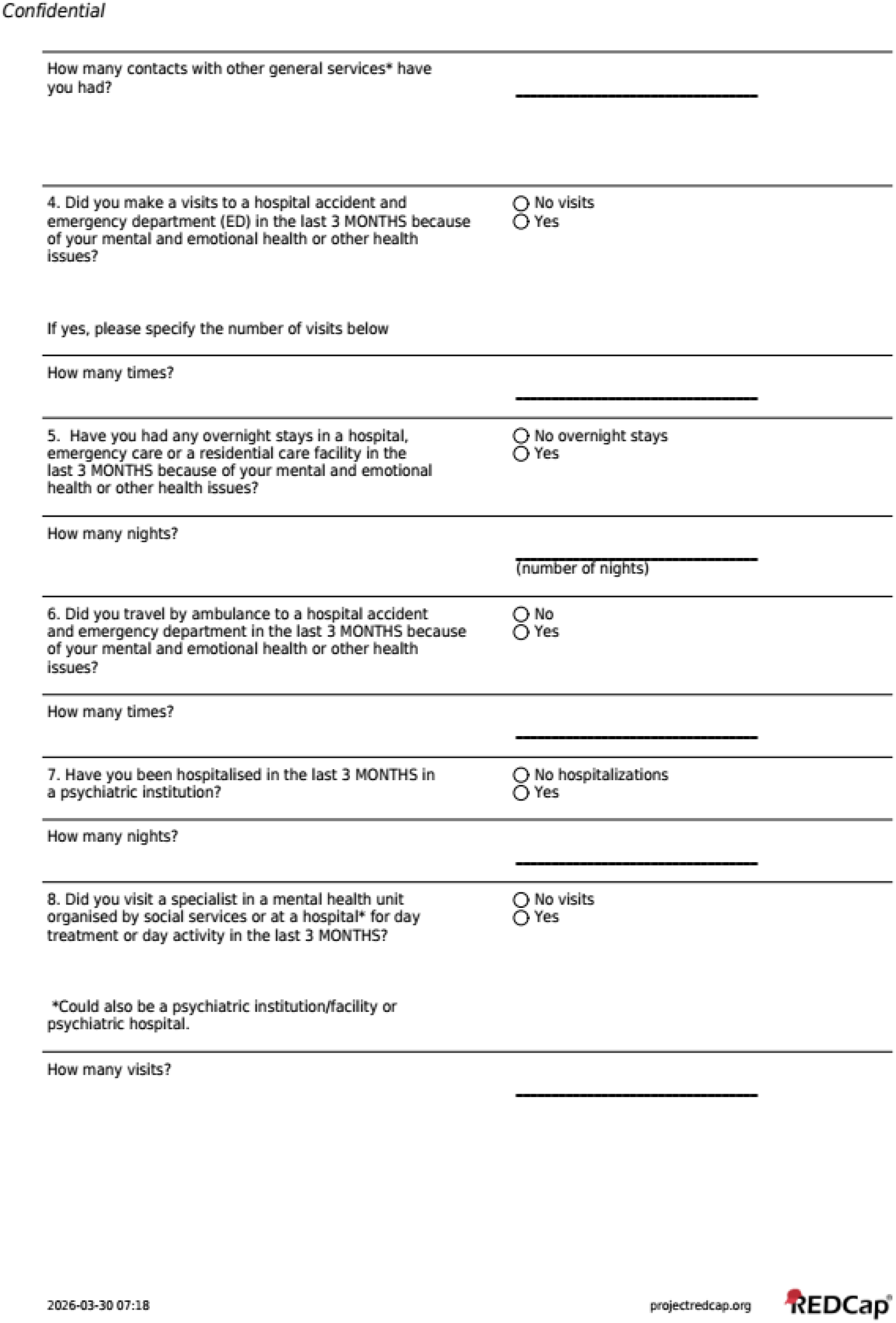
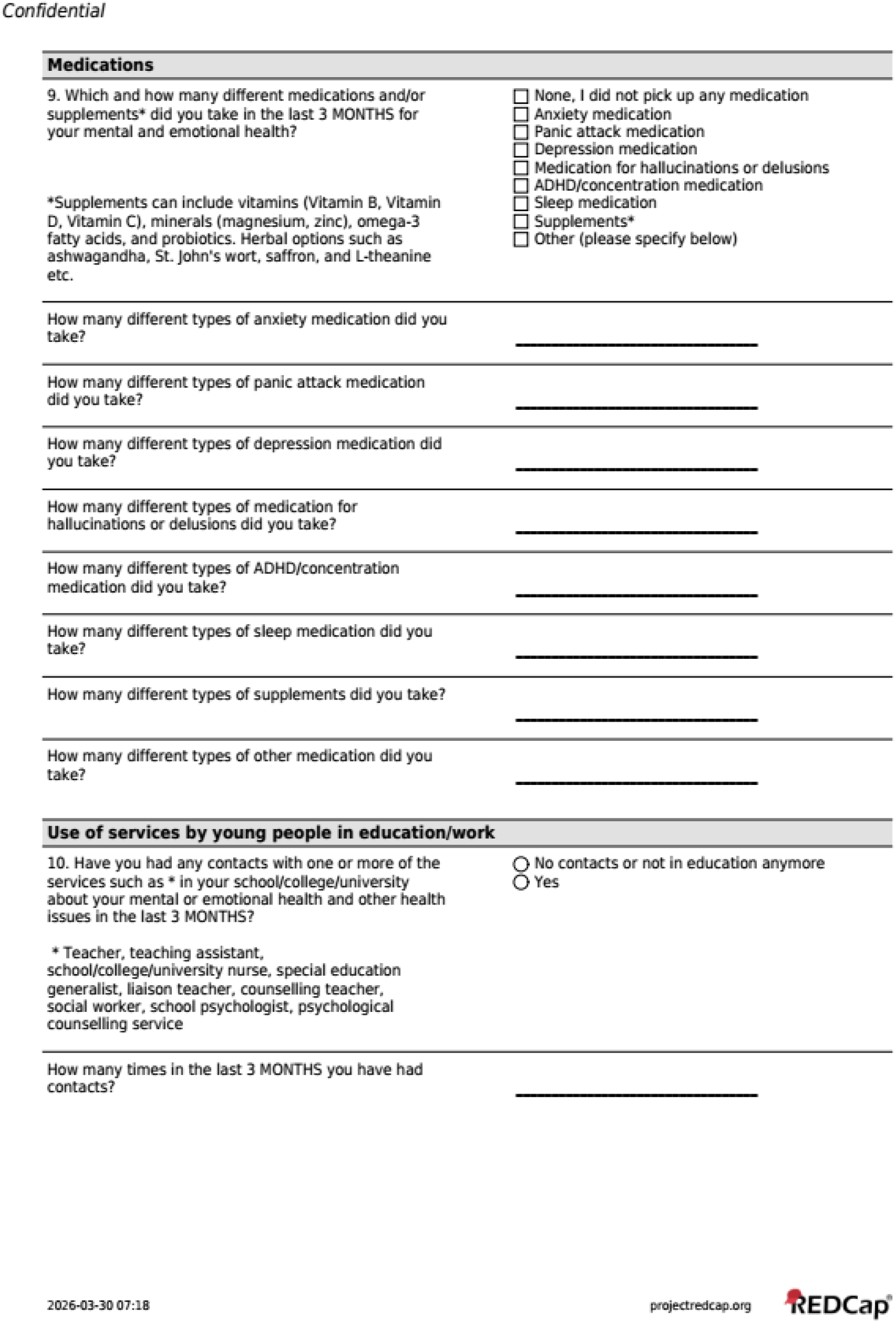
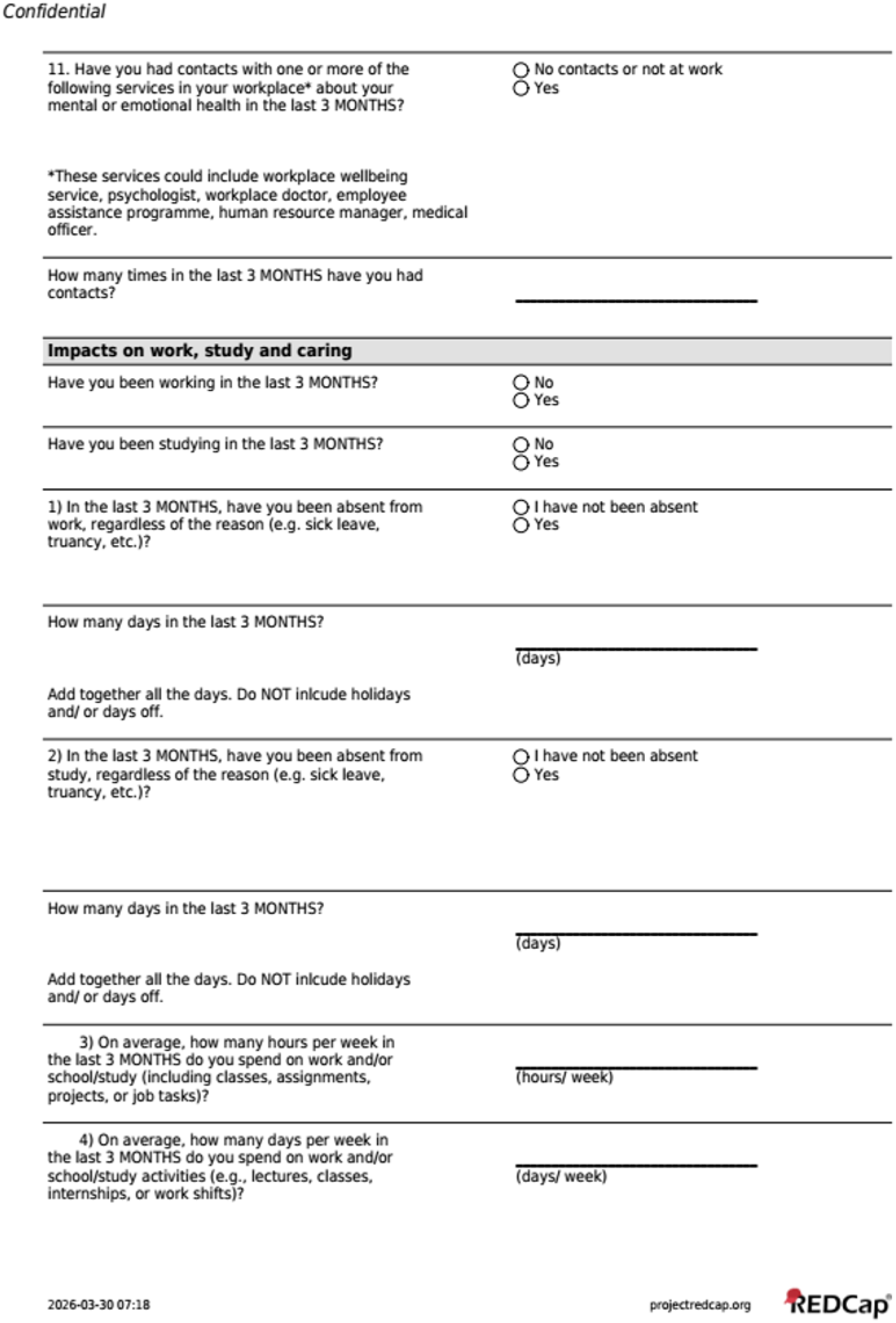
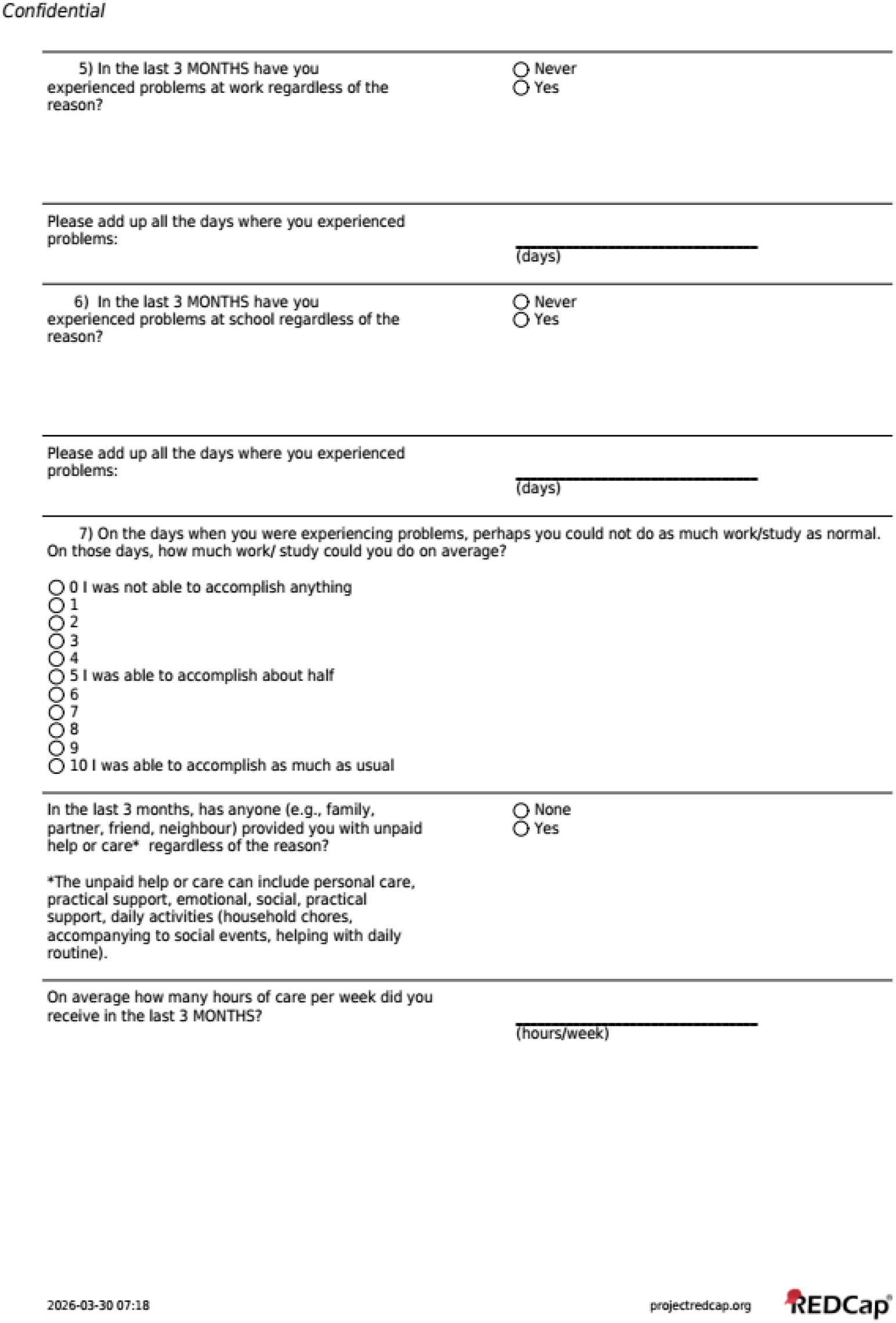

### 2.3 Resource Use Measurement Survey for the Netherlands

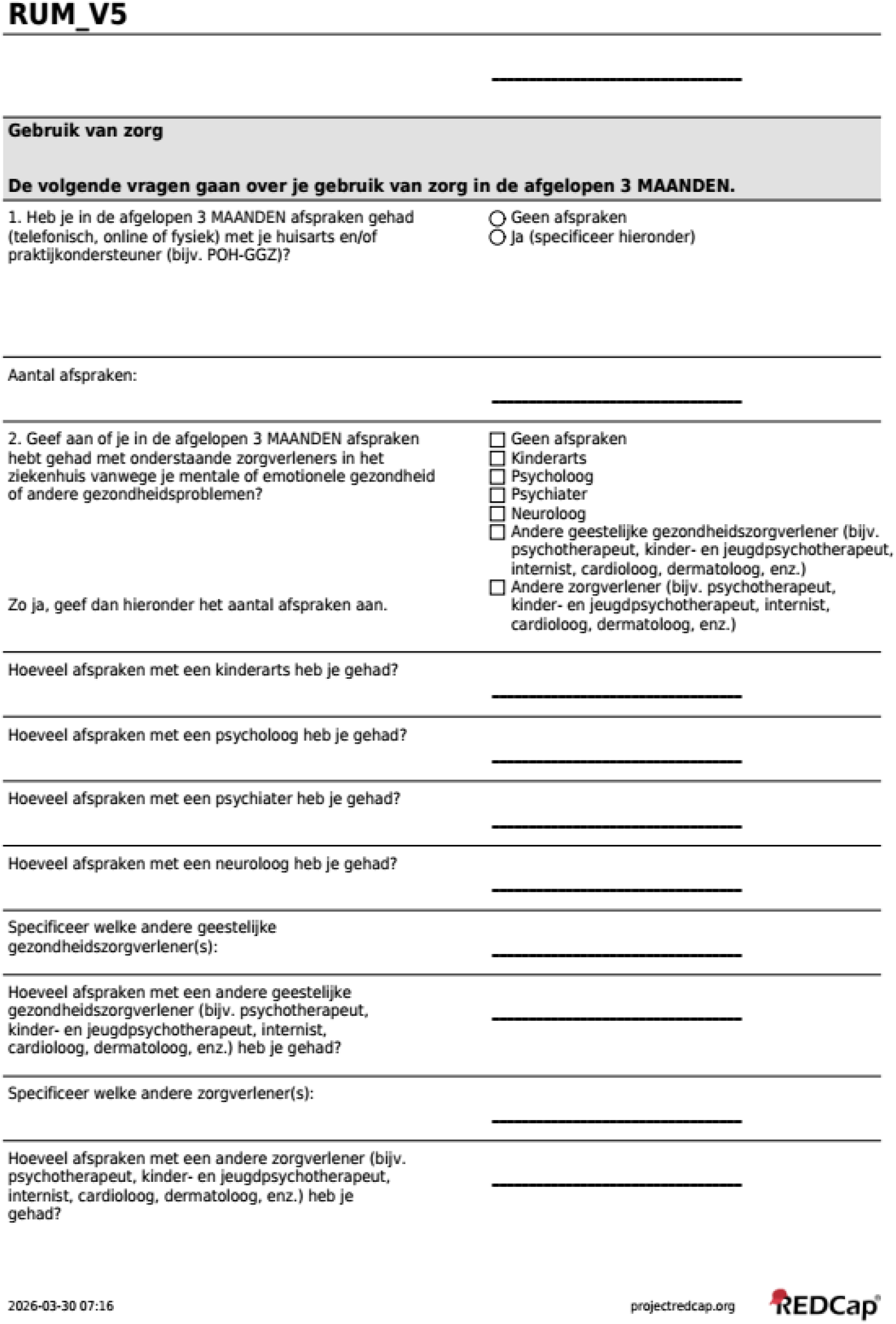
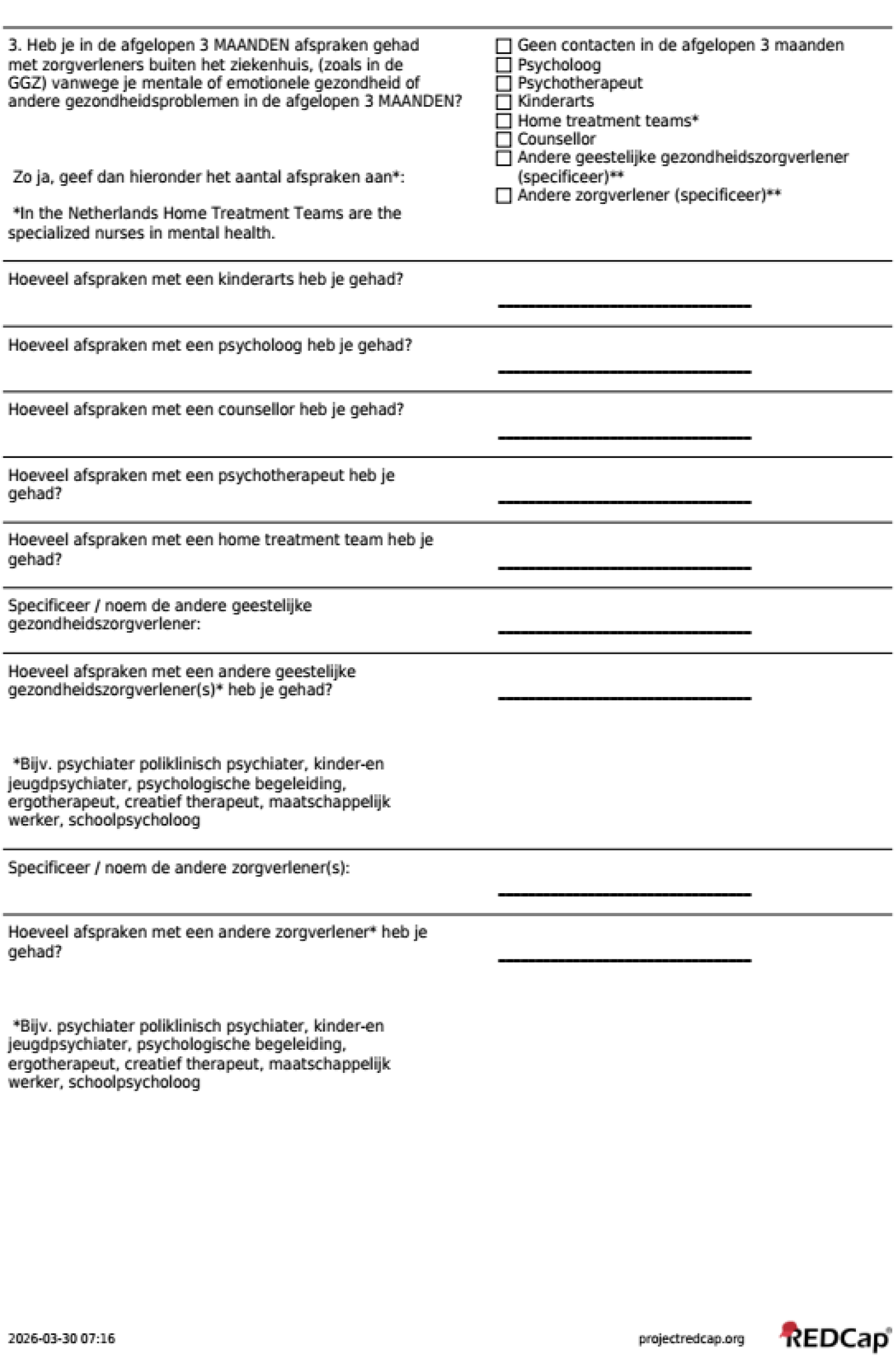
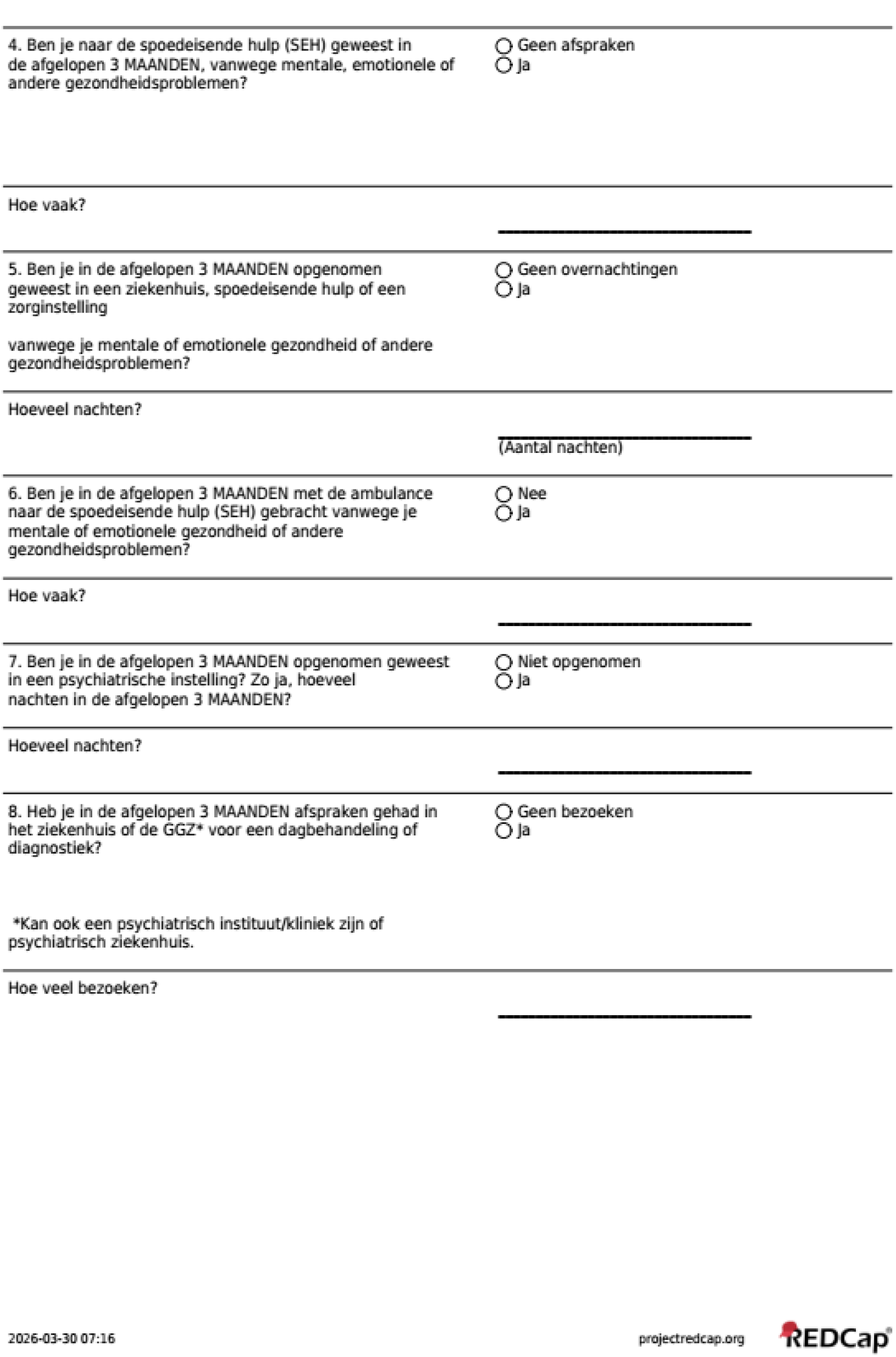
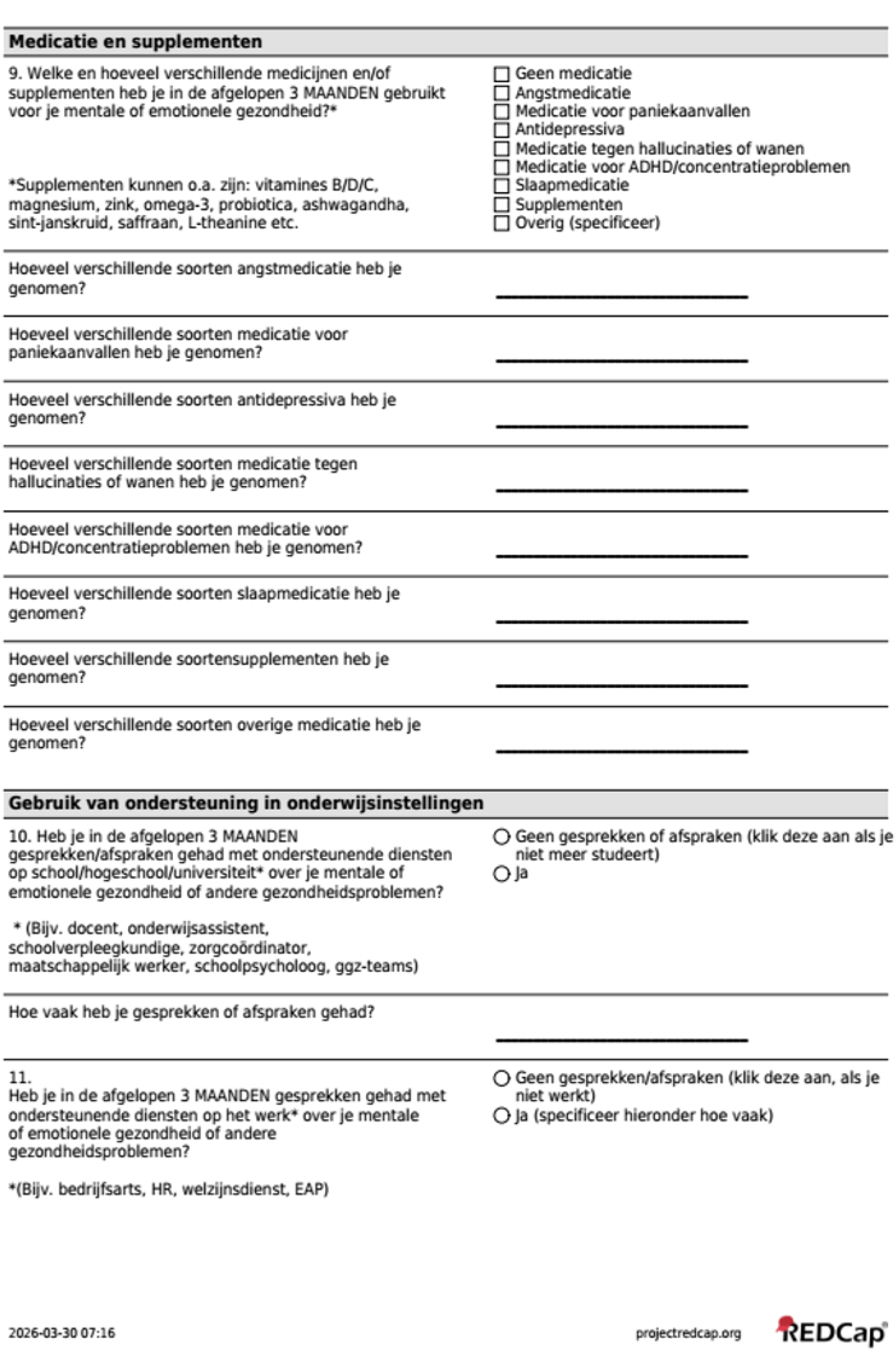
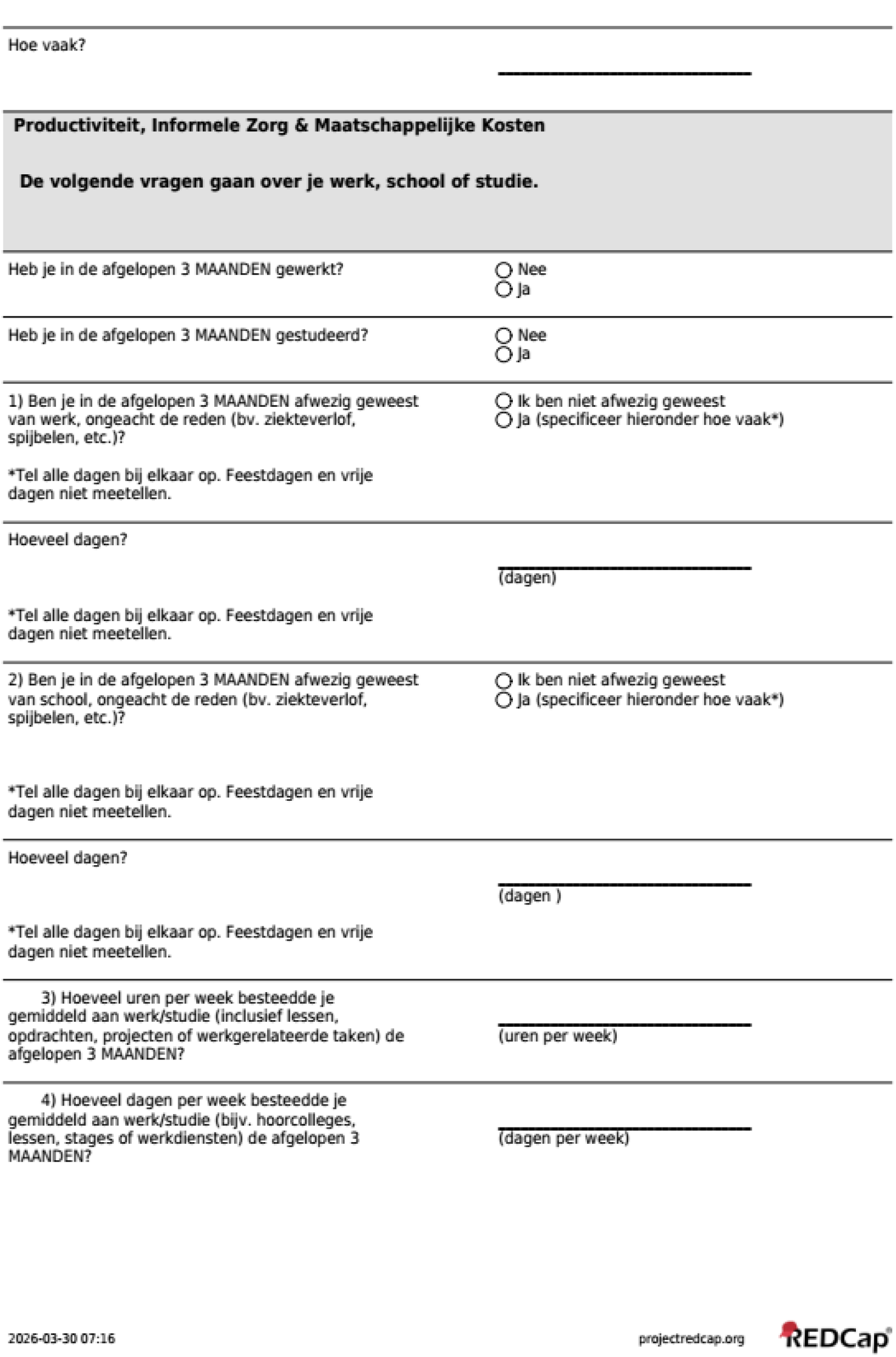
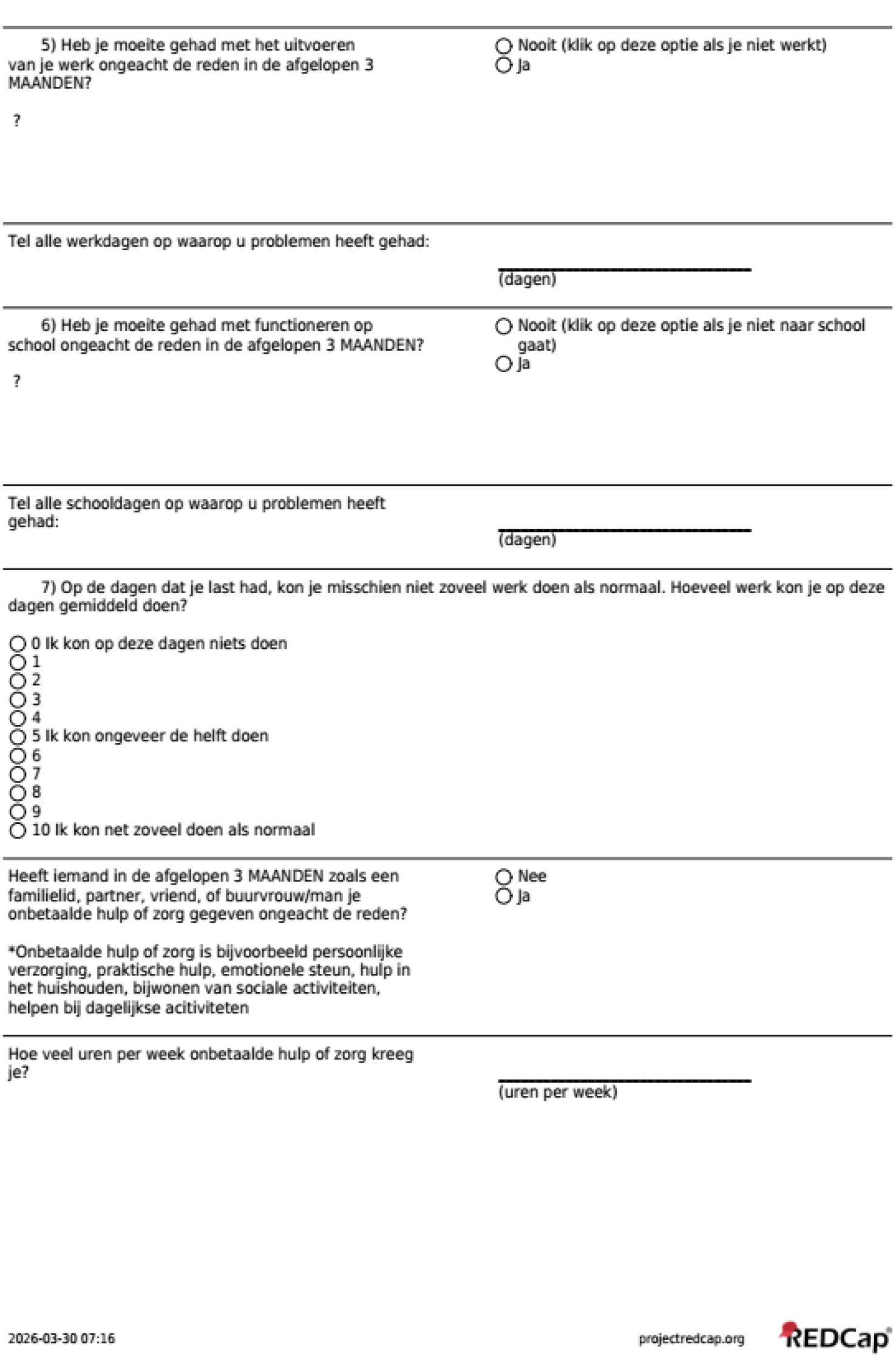

### 2.4 Resource Use Measurement Survey for Germany

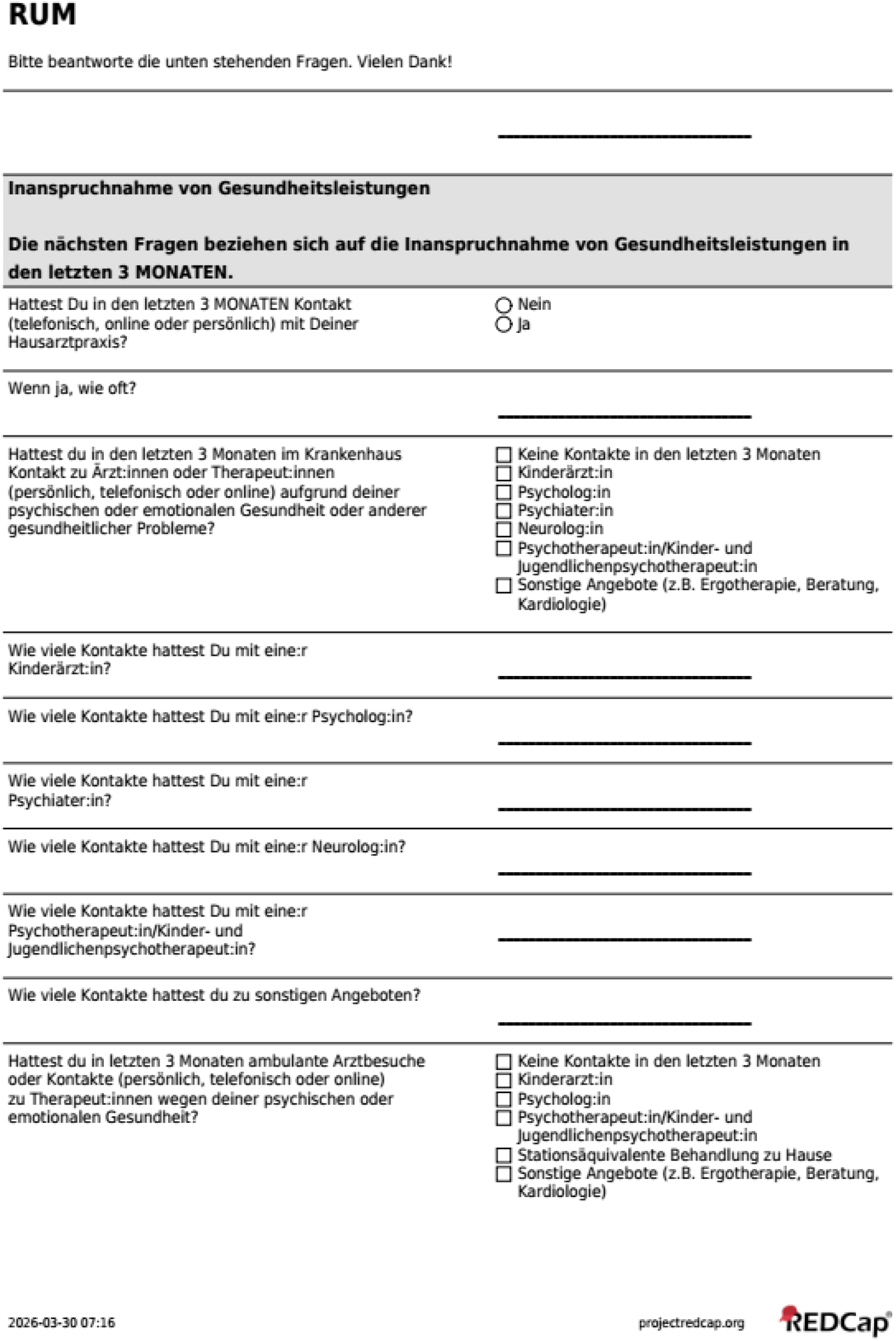
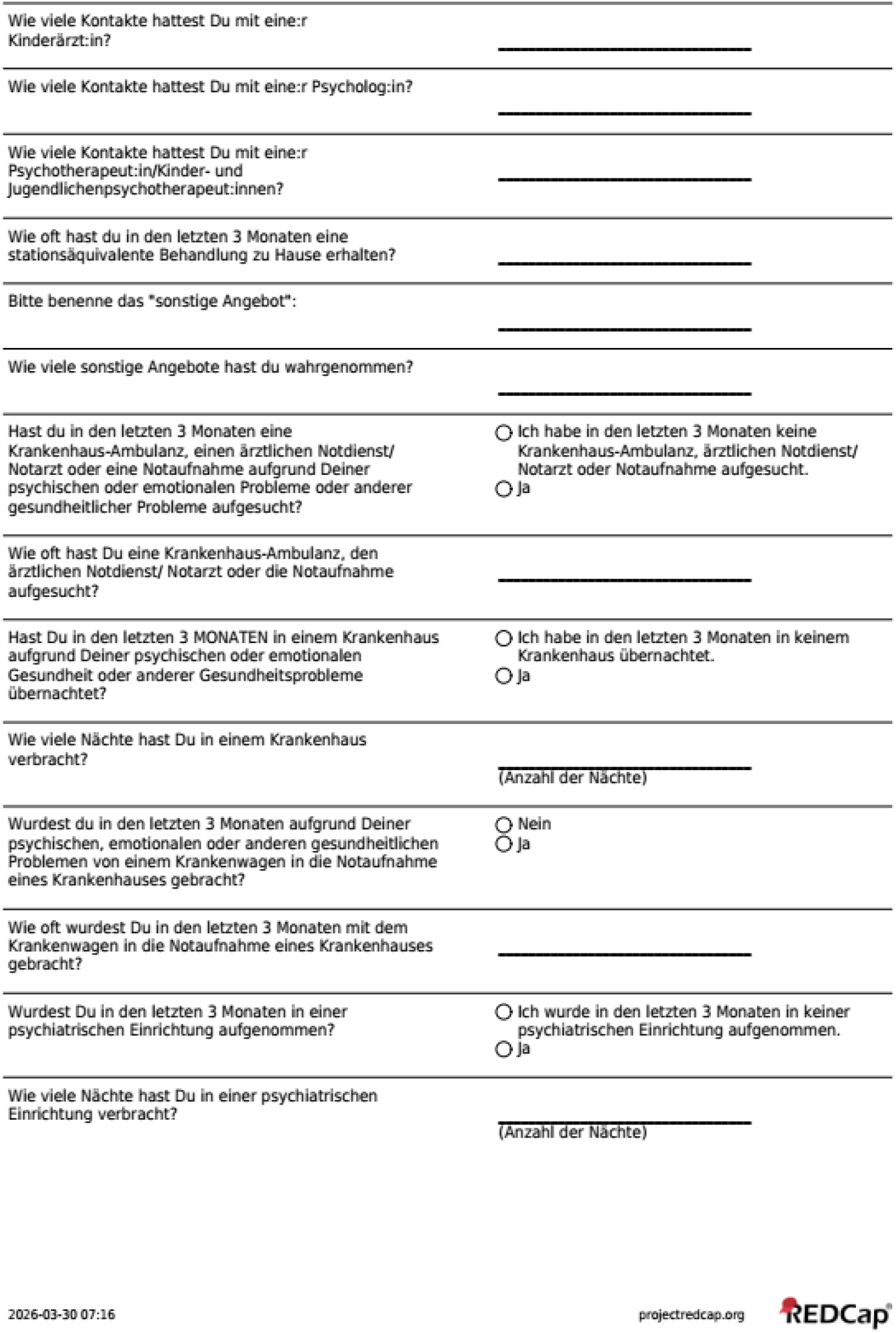
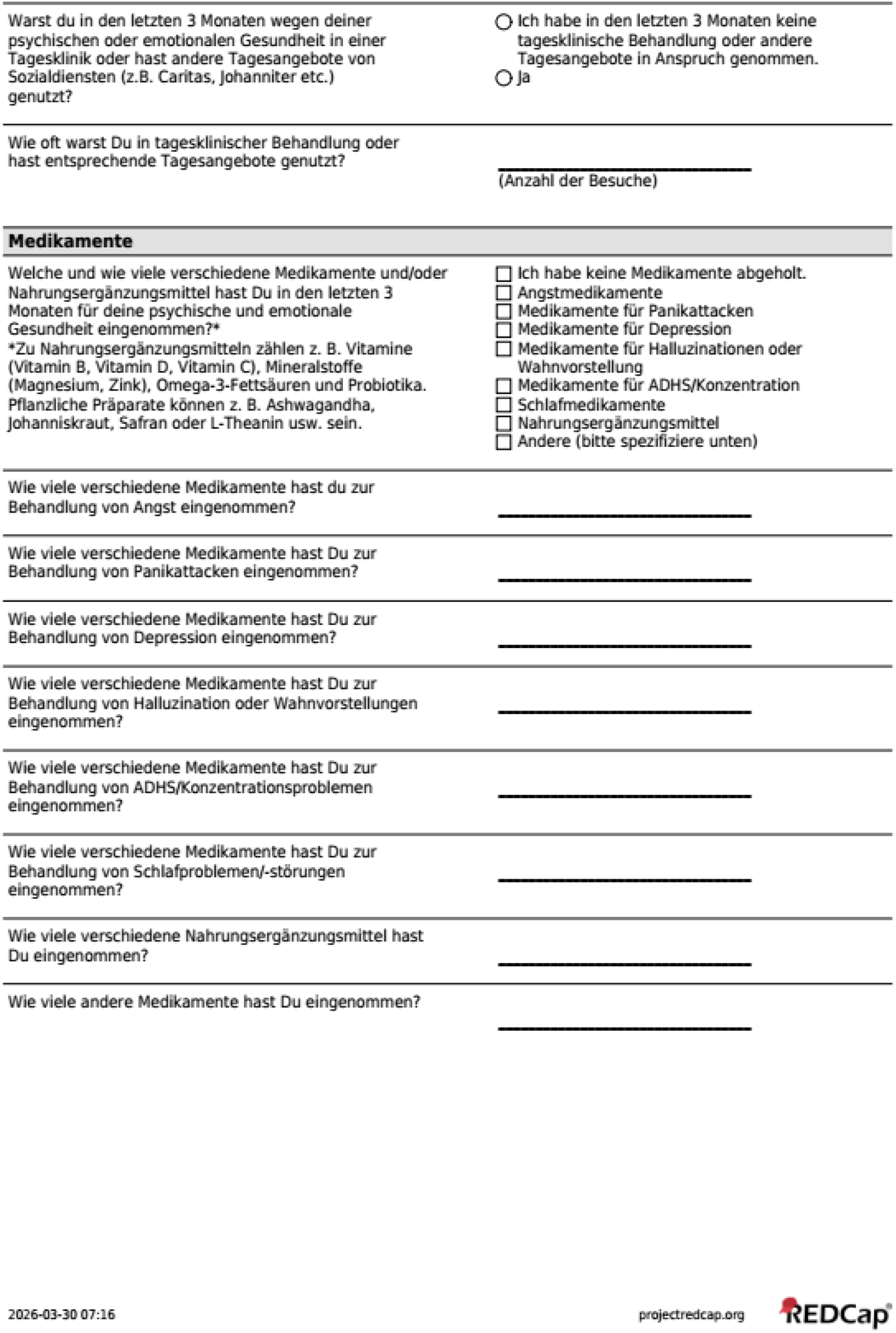
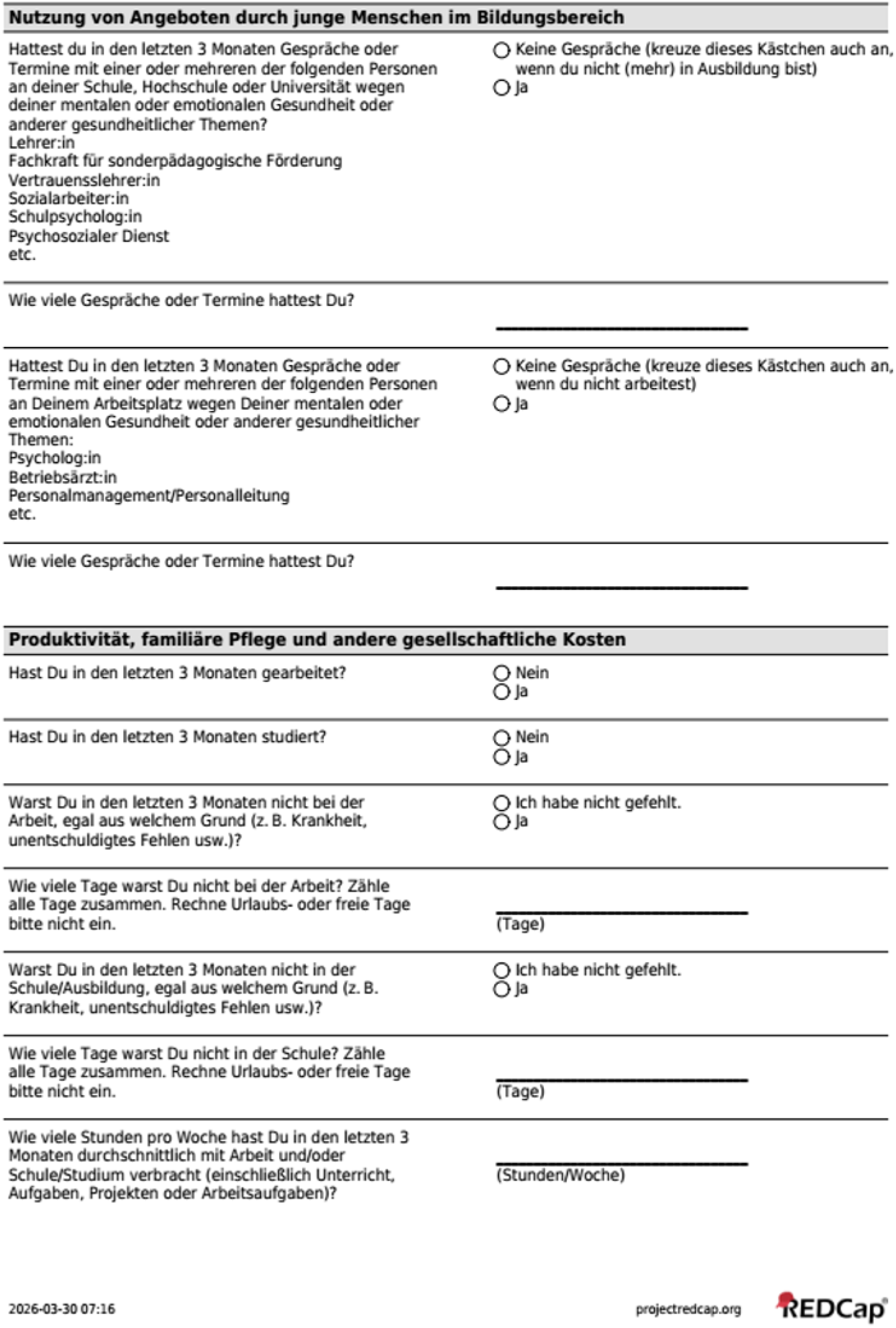
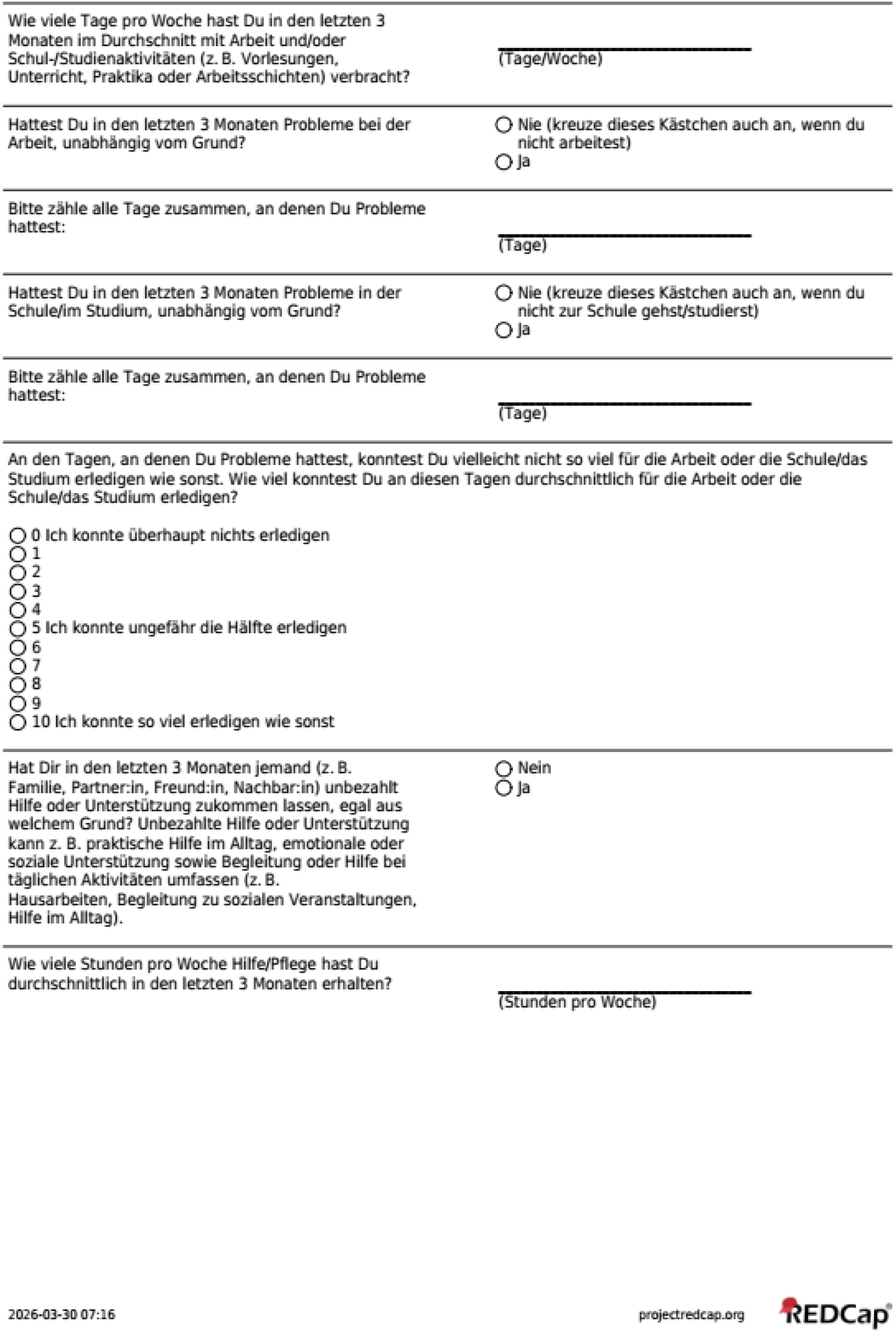

### 2.5 Resource Use Measurement Survey for Italy

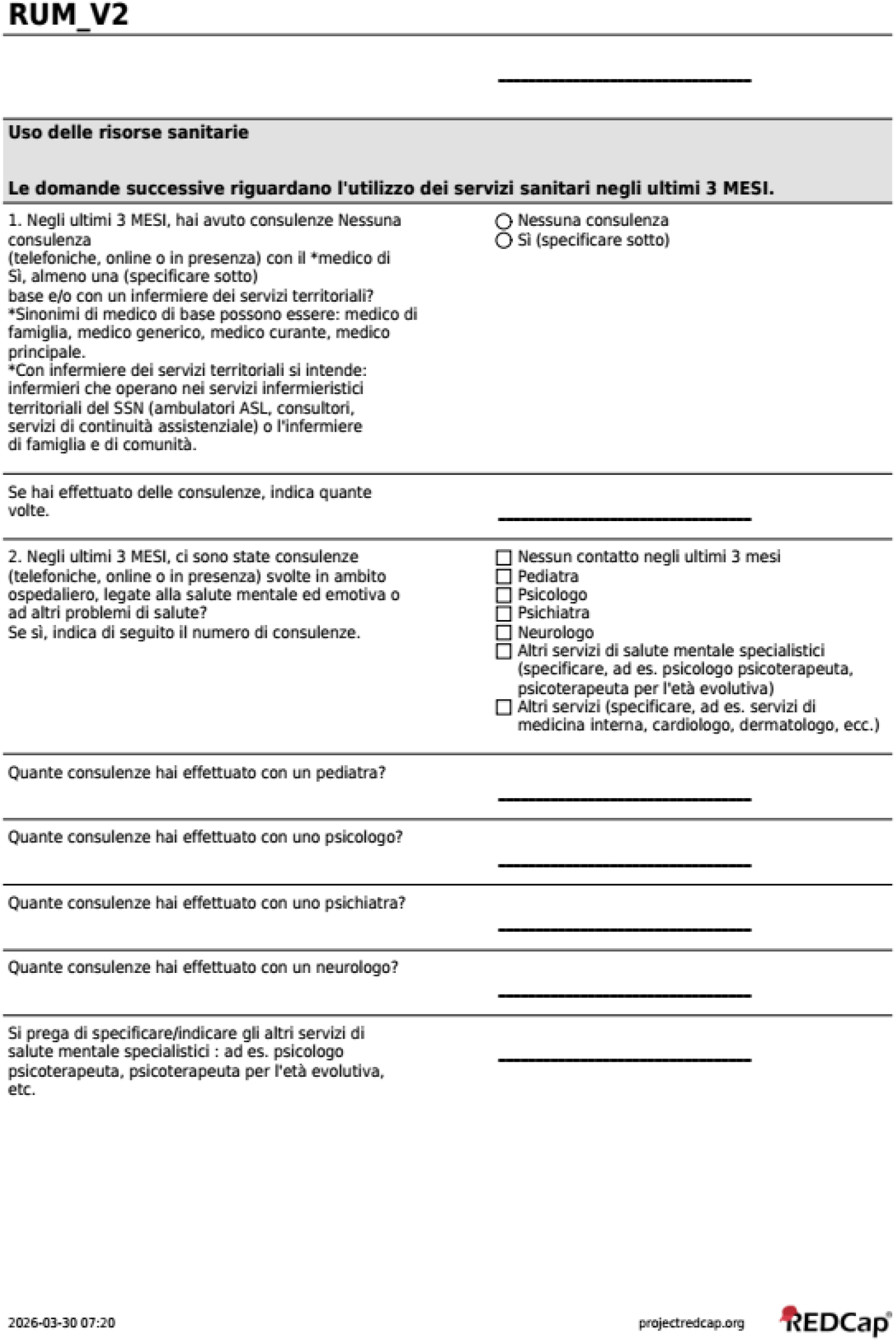
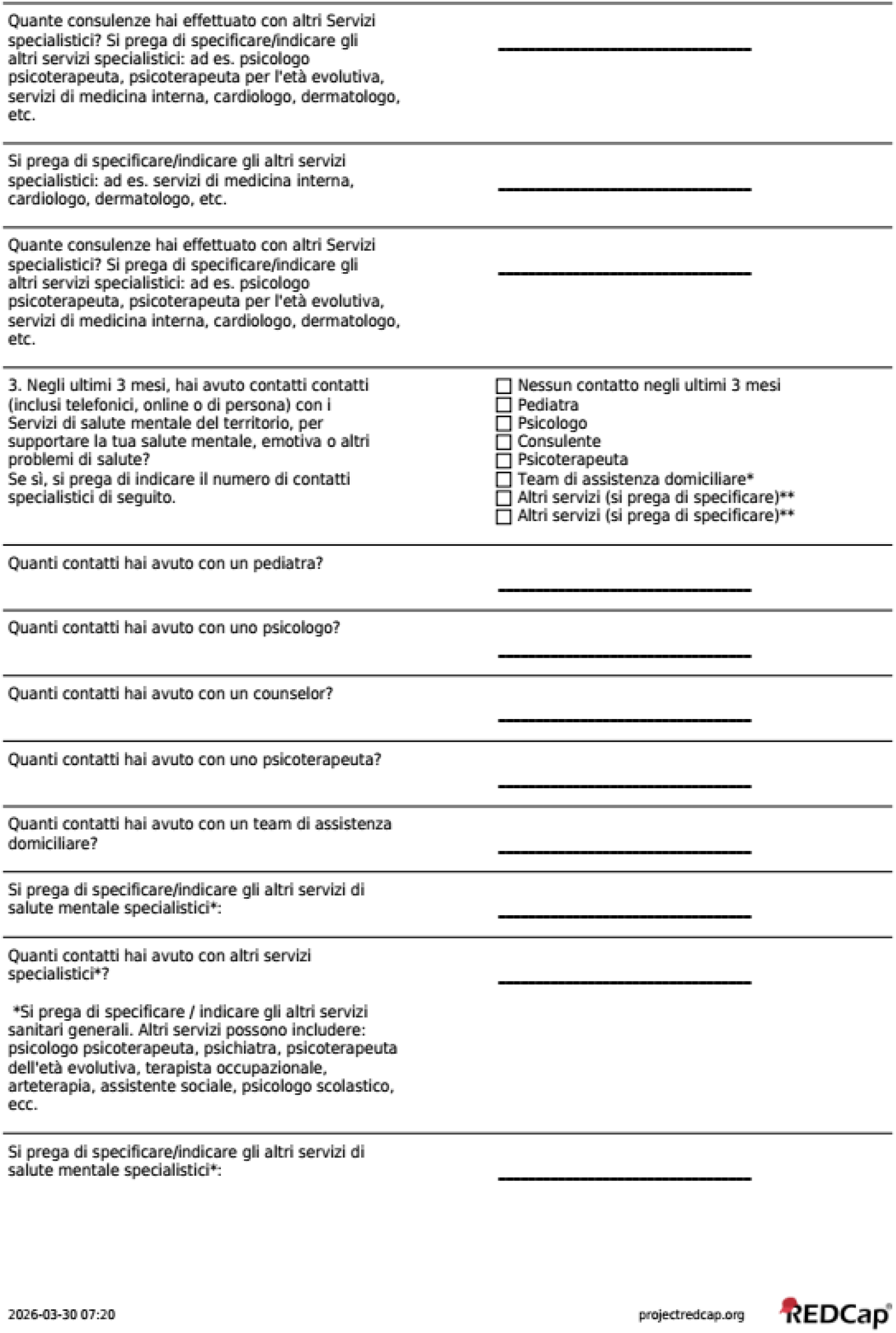
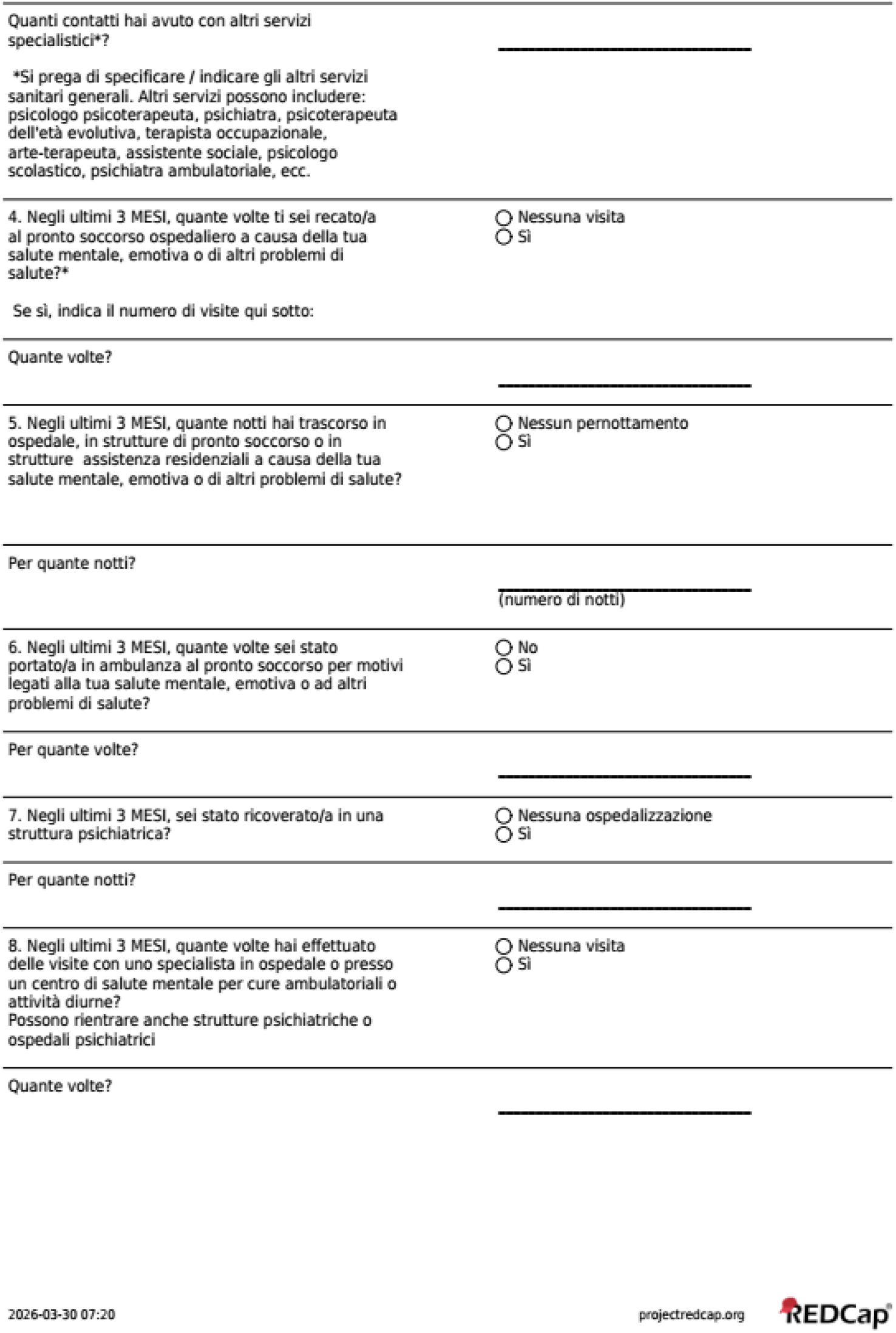
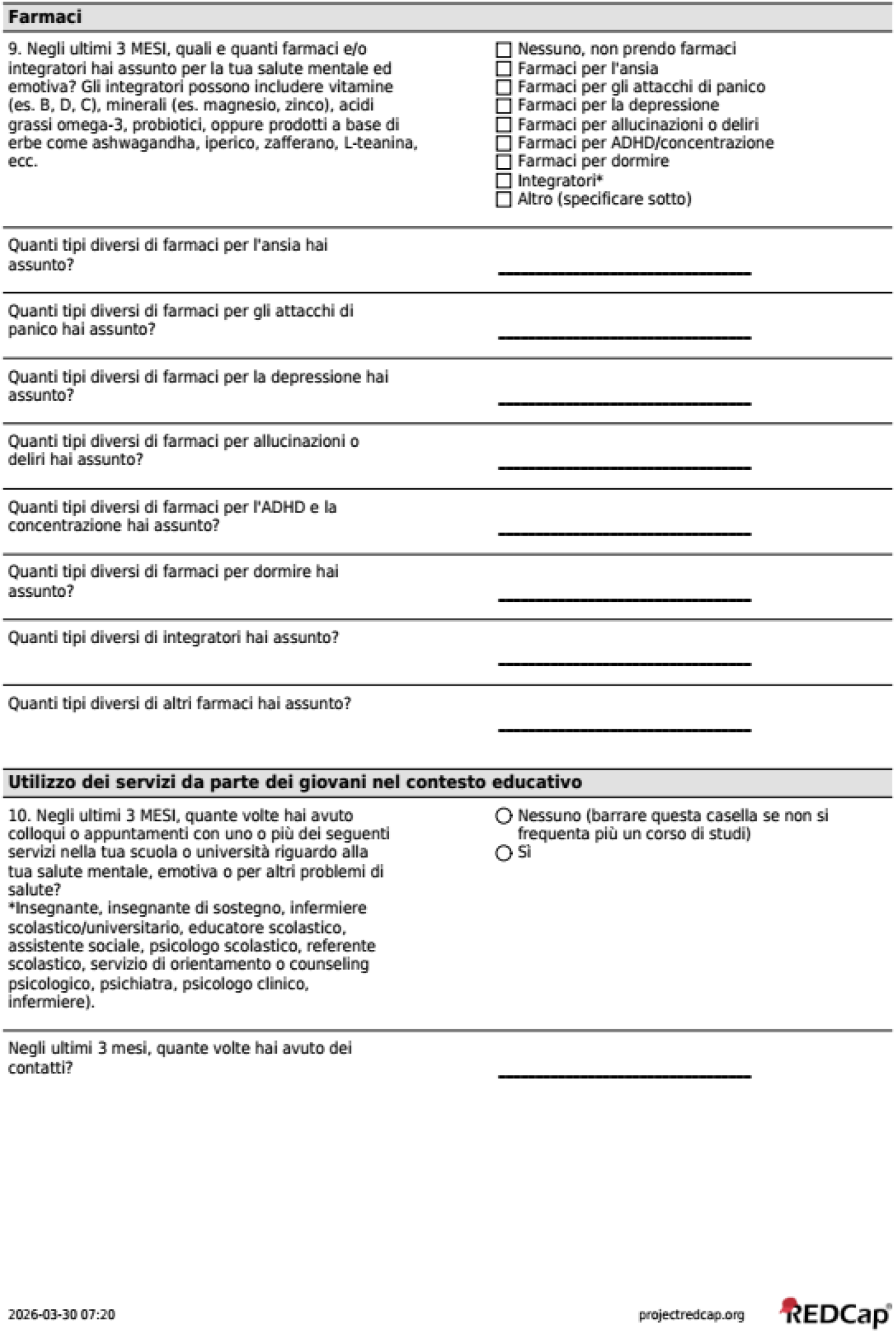
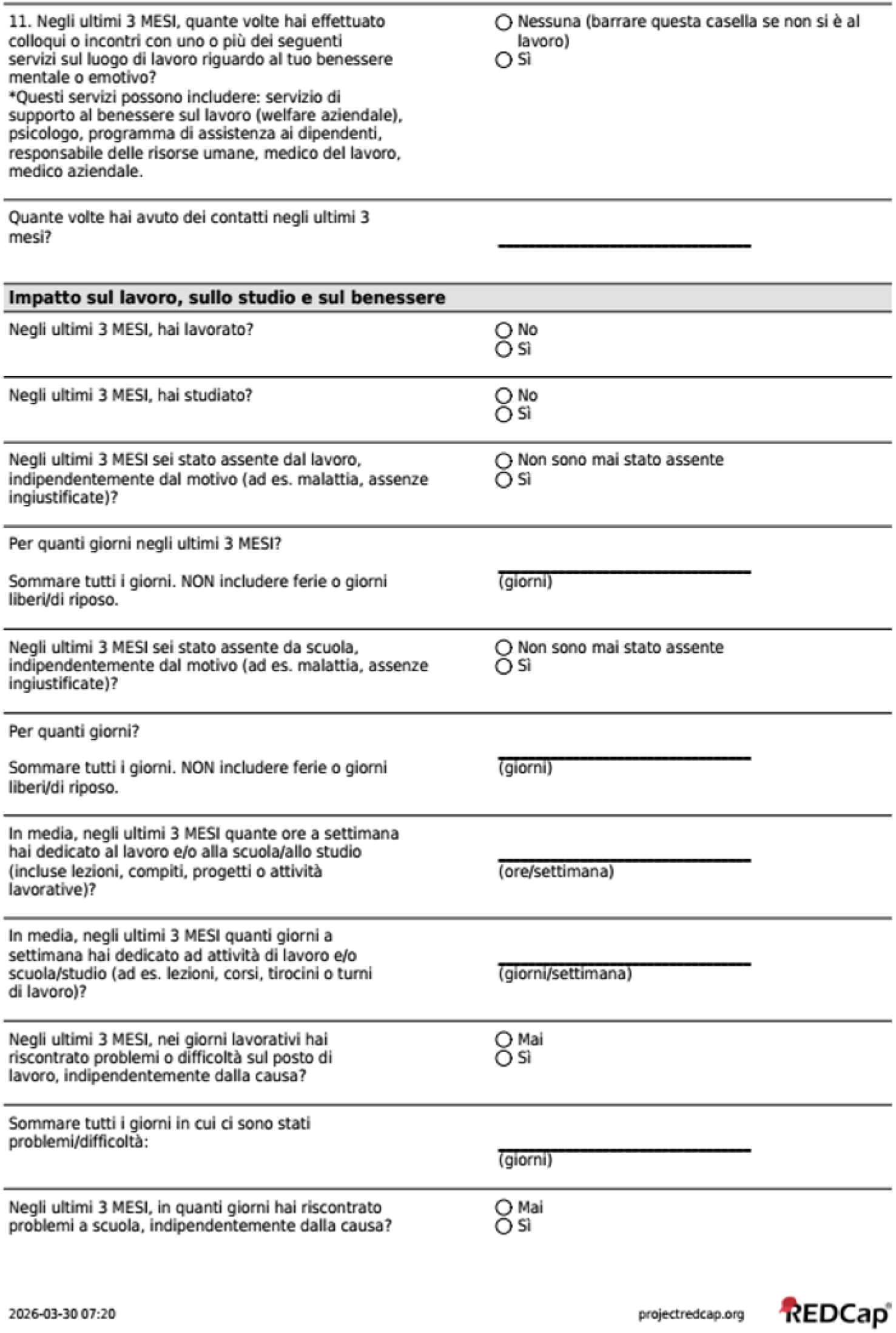
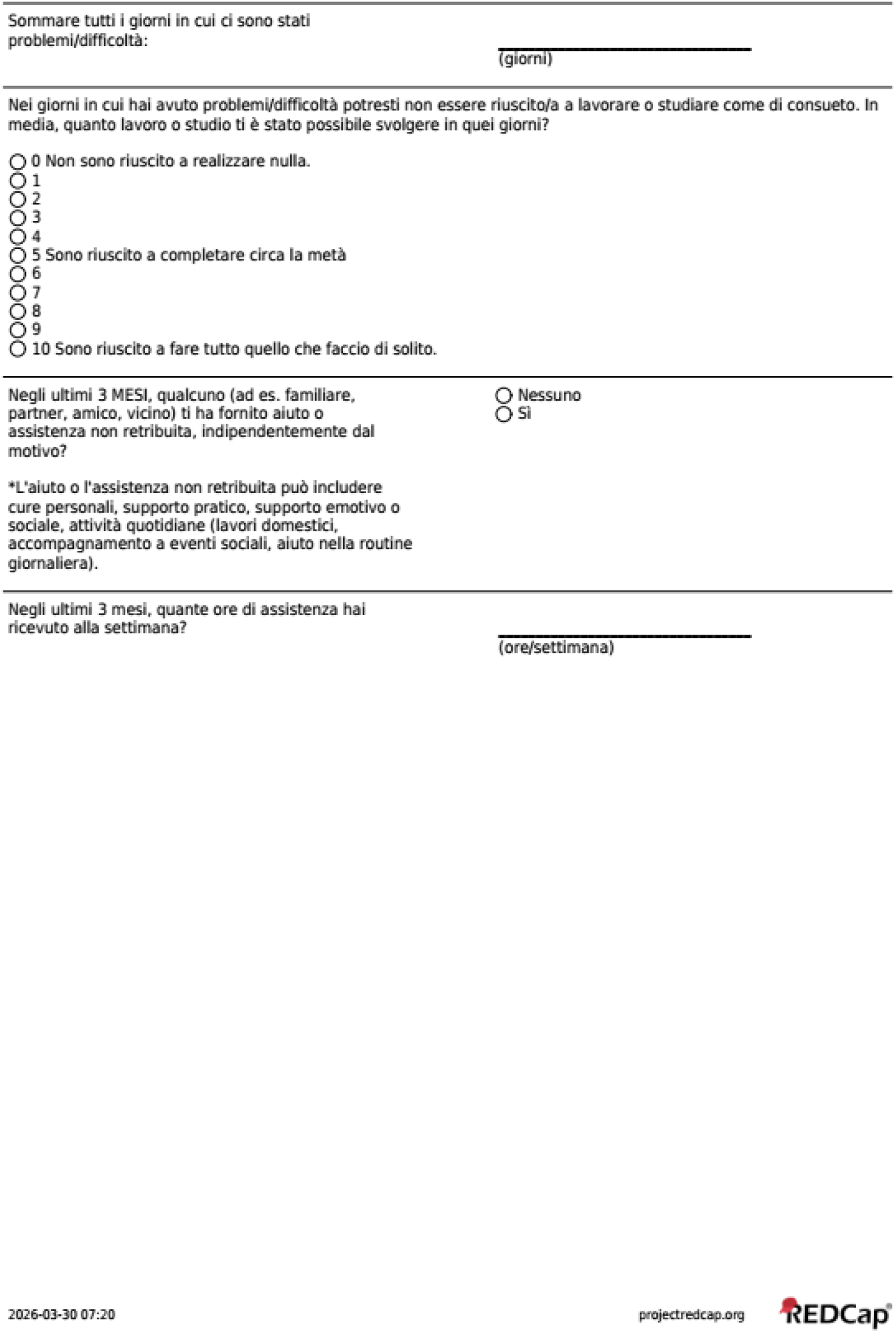

### 2.6 Resource Use Measurement Survey for Spain

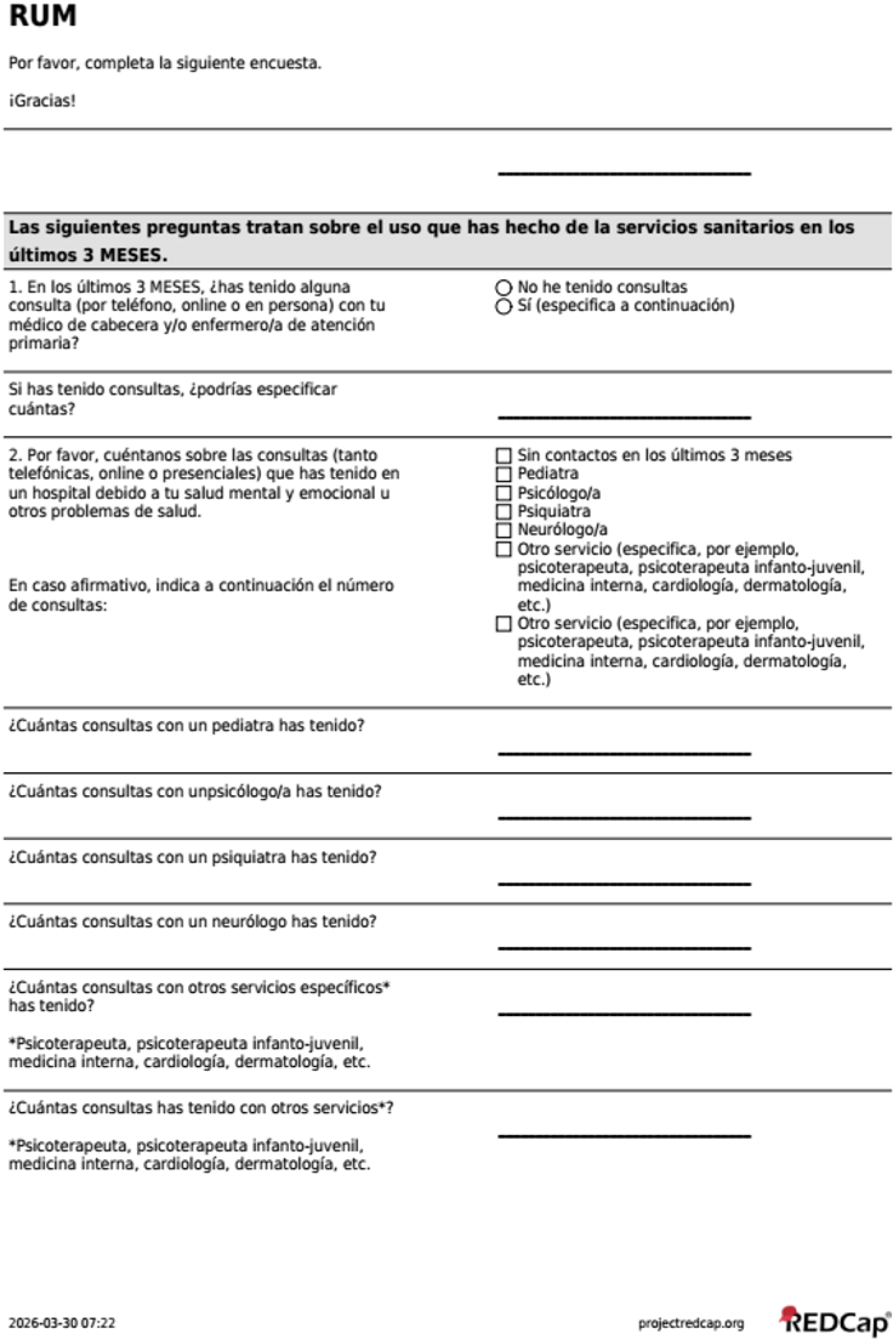
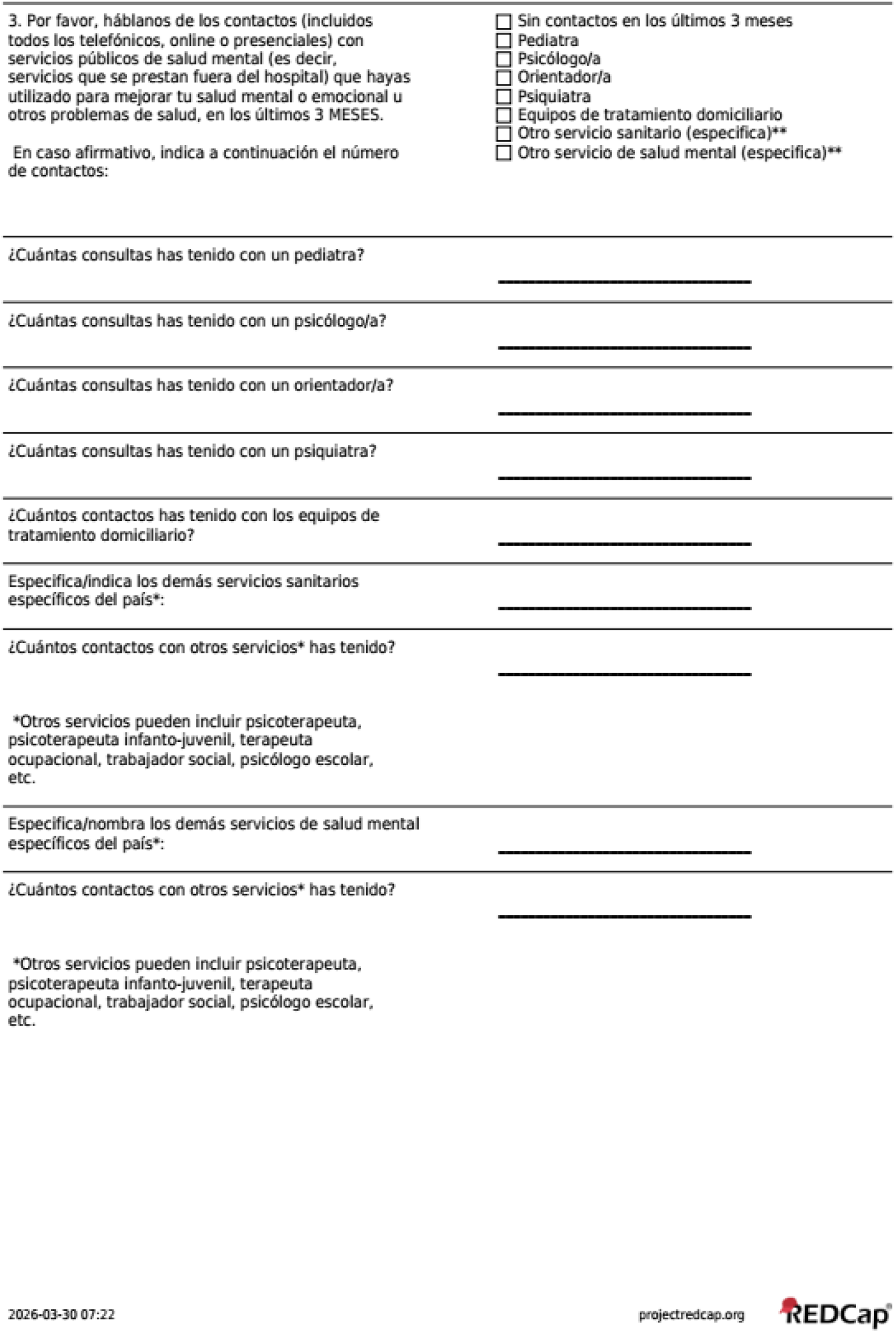
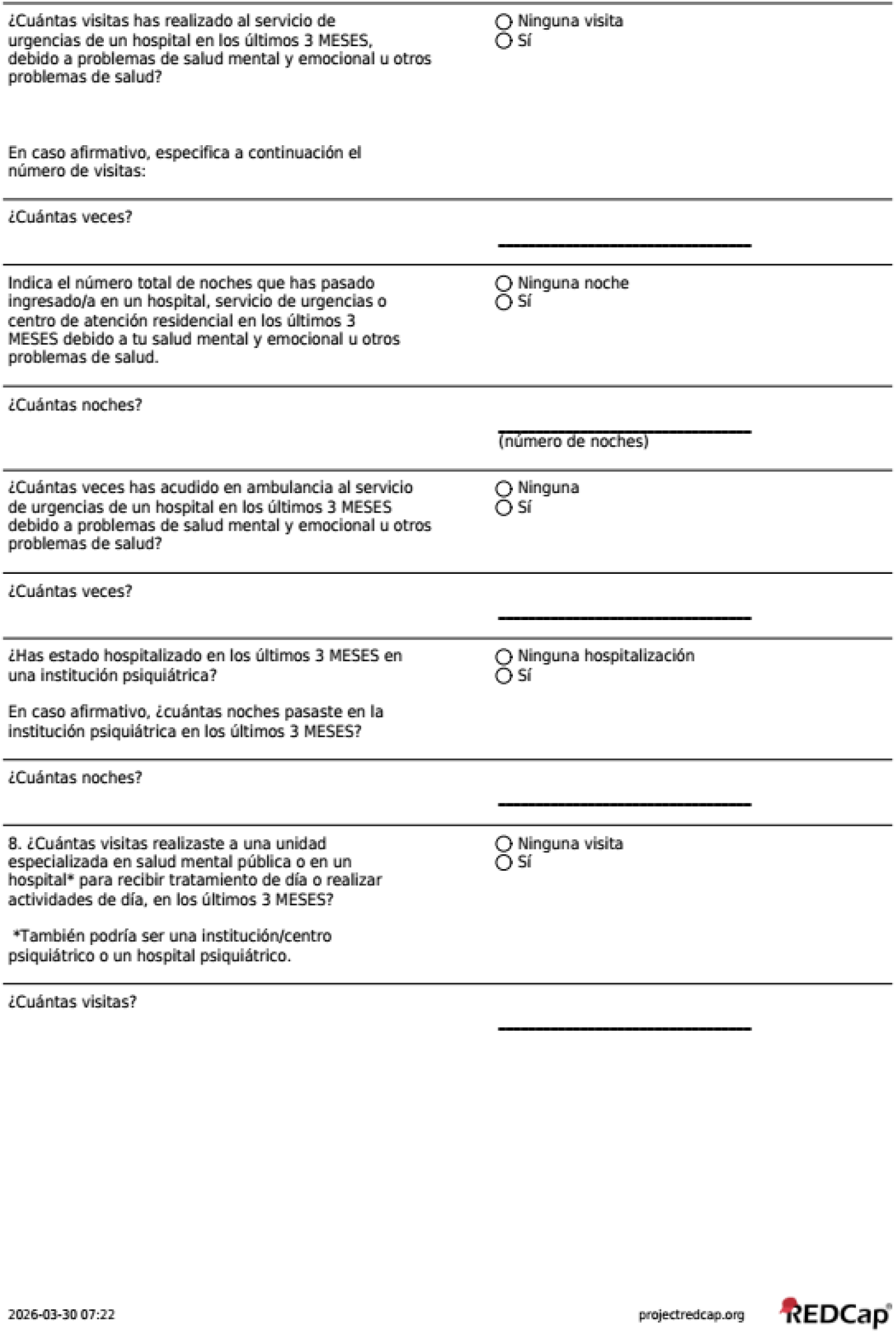
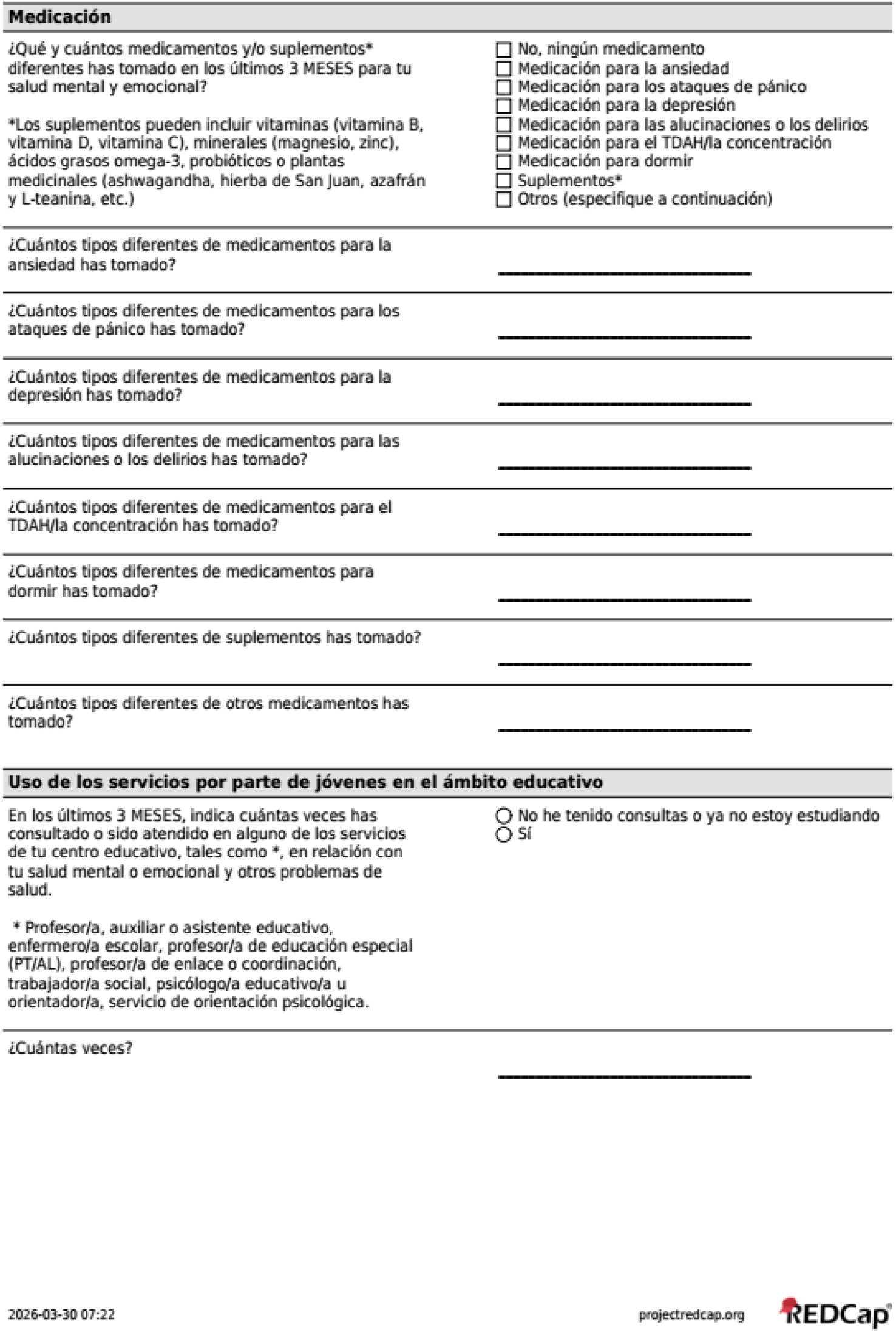
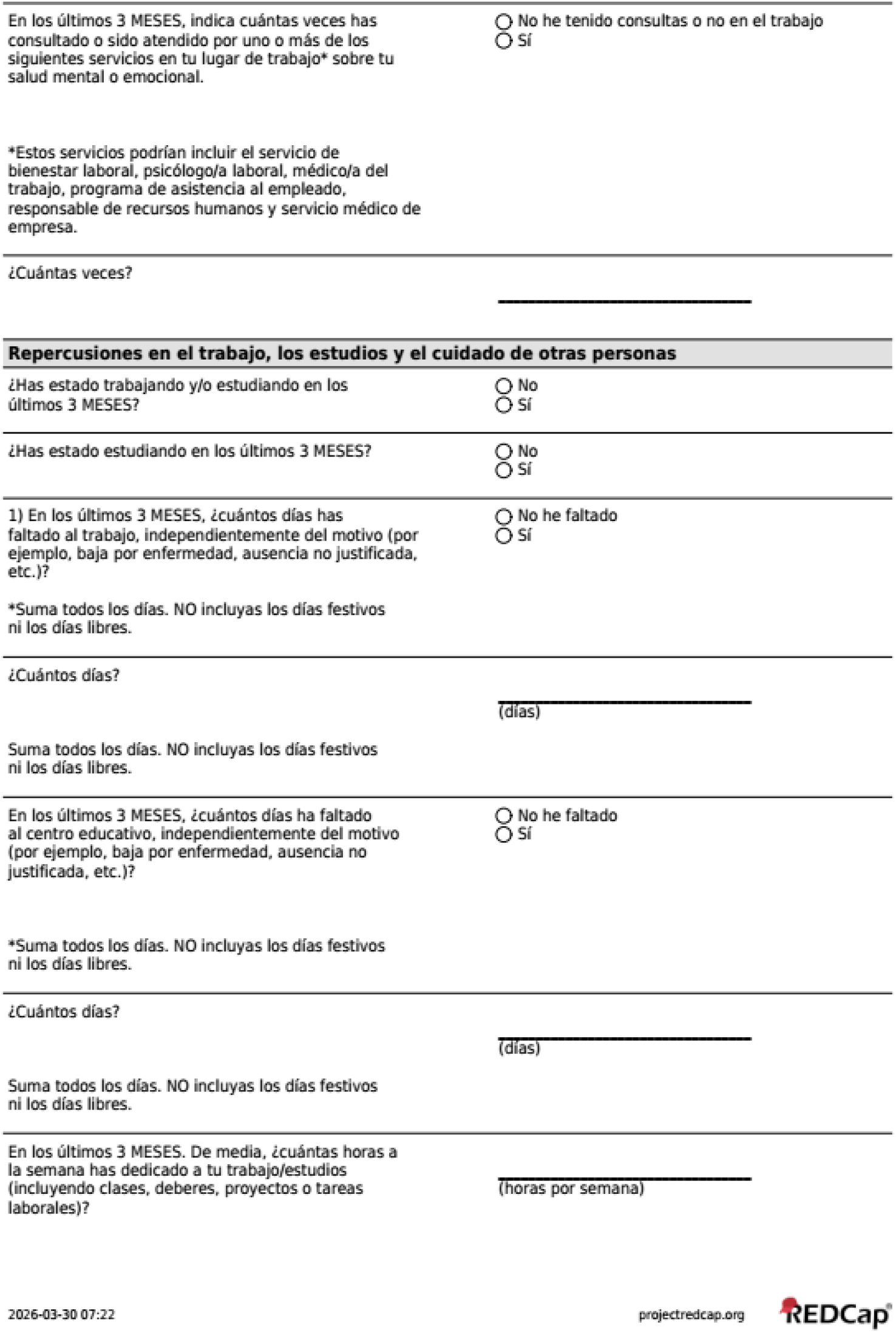
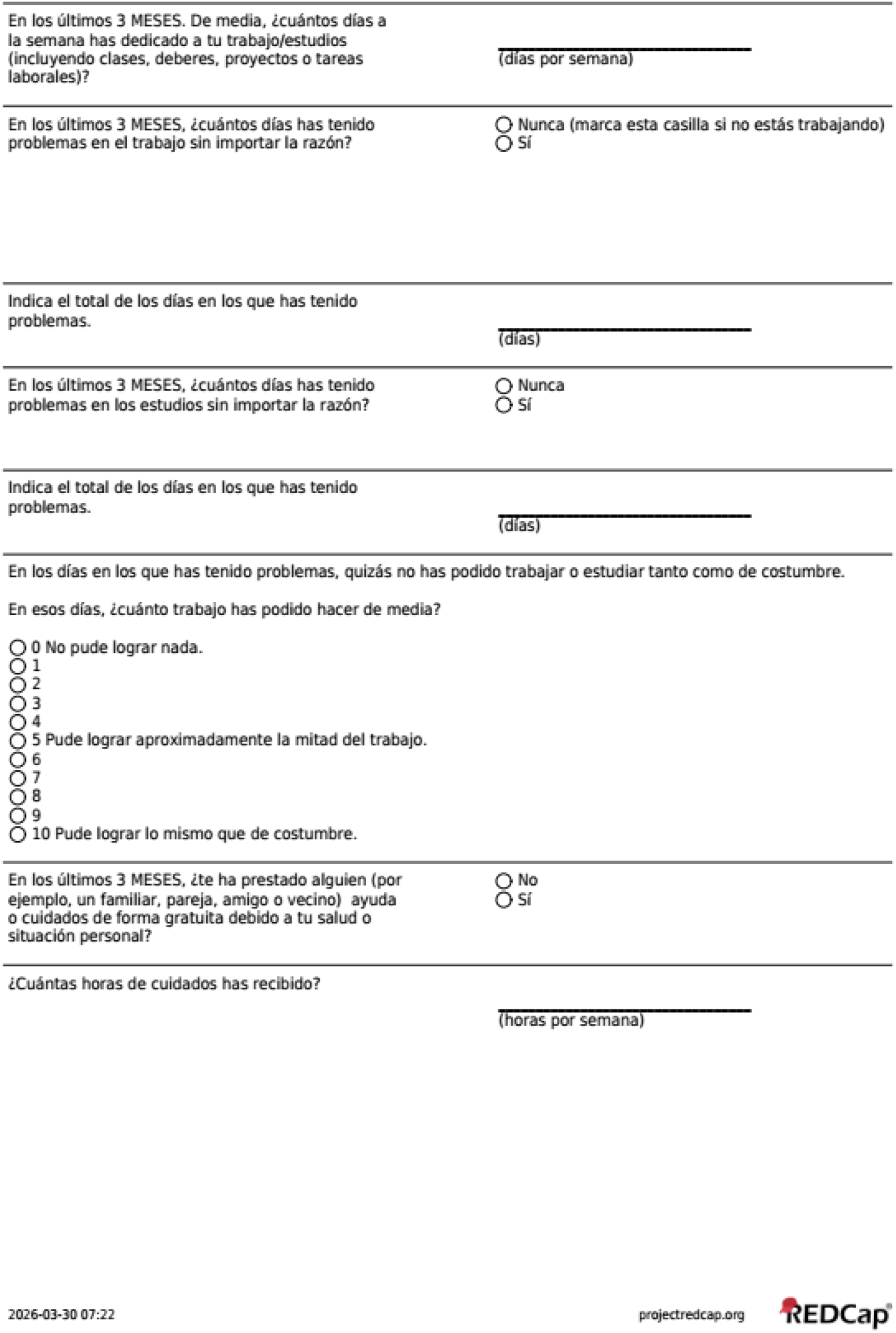

### 2.7 Resource Use Measurement Survey for Estonia

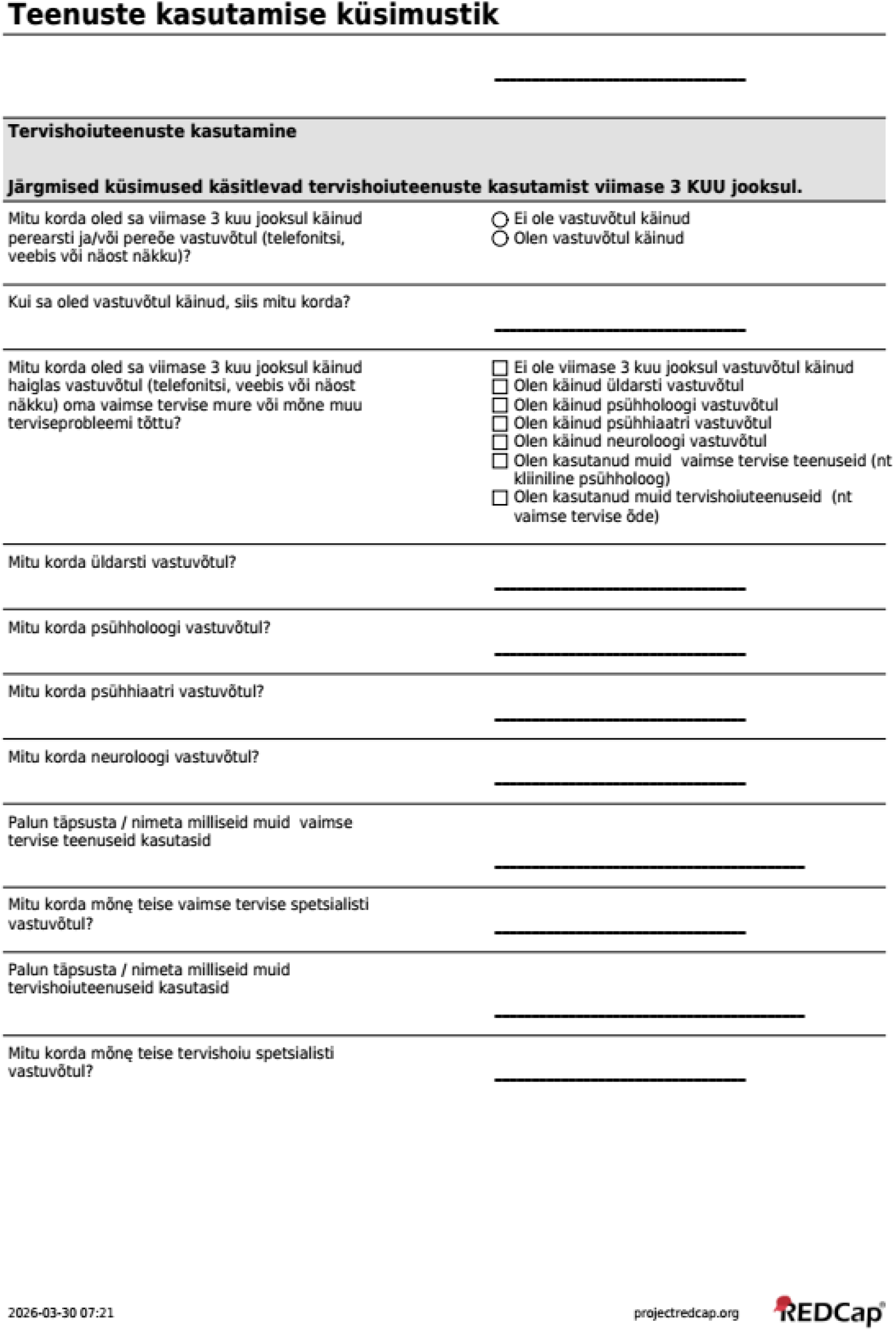
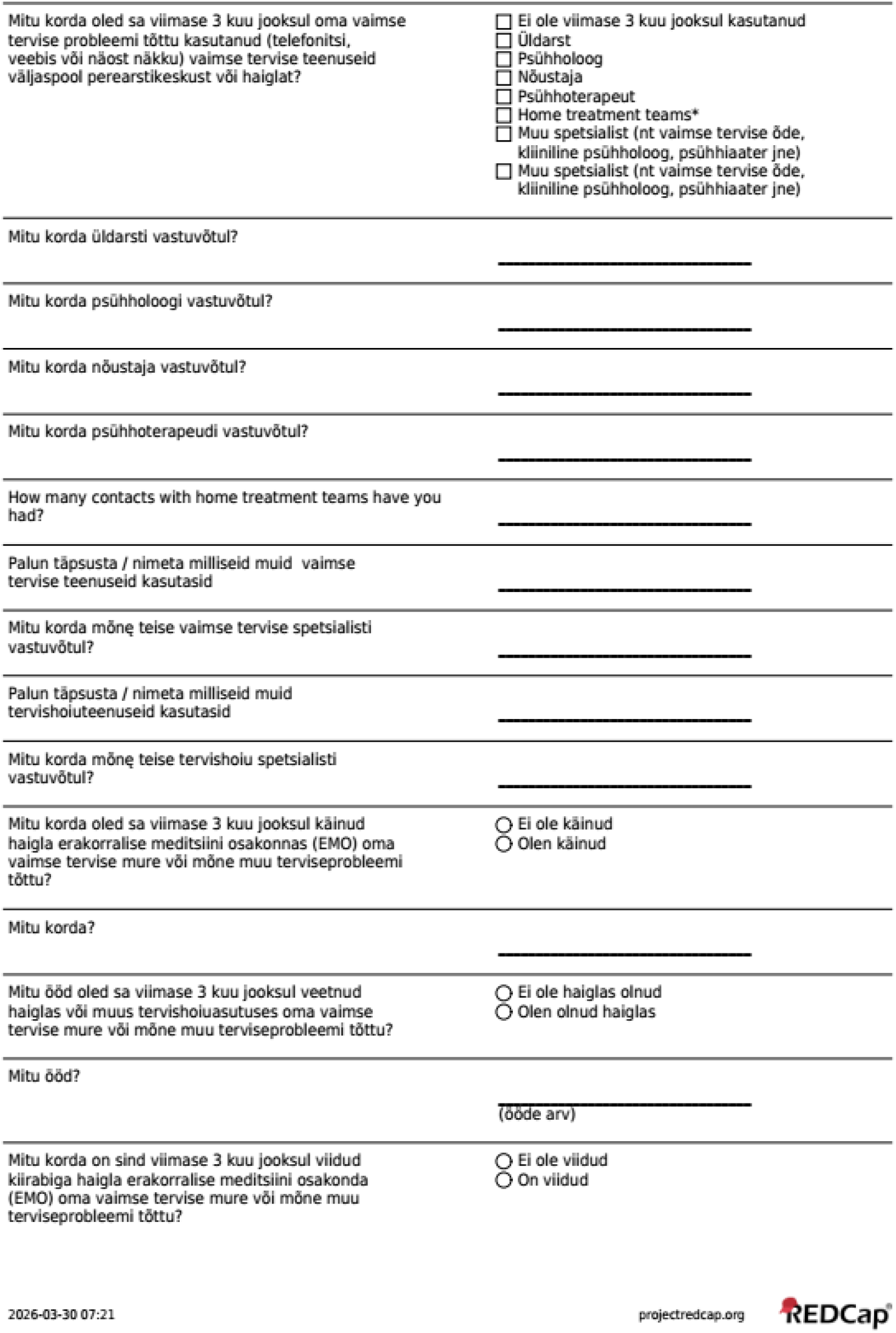
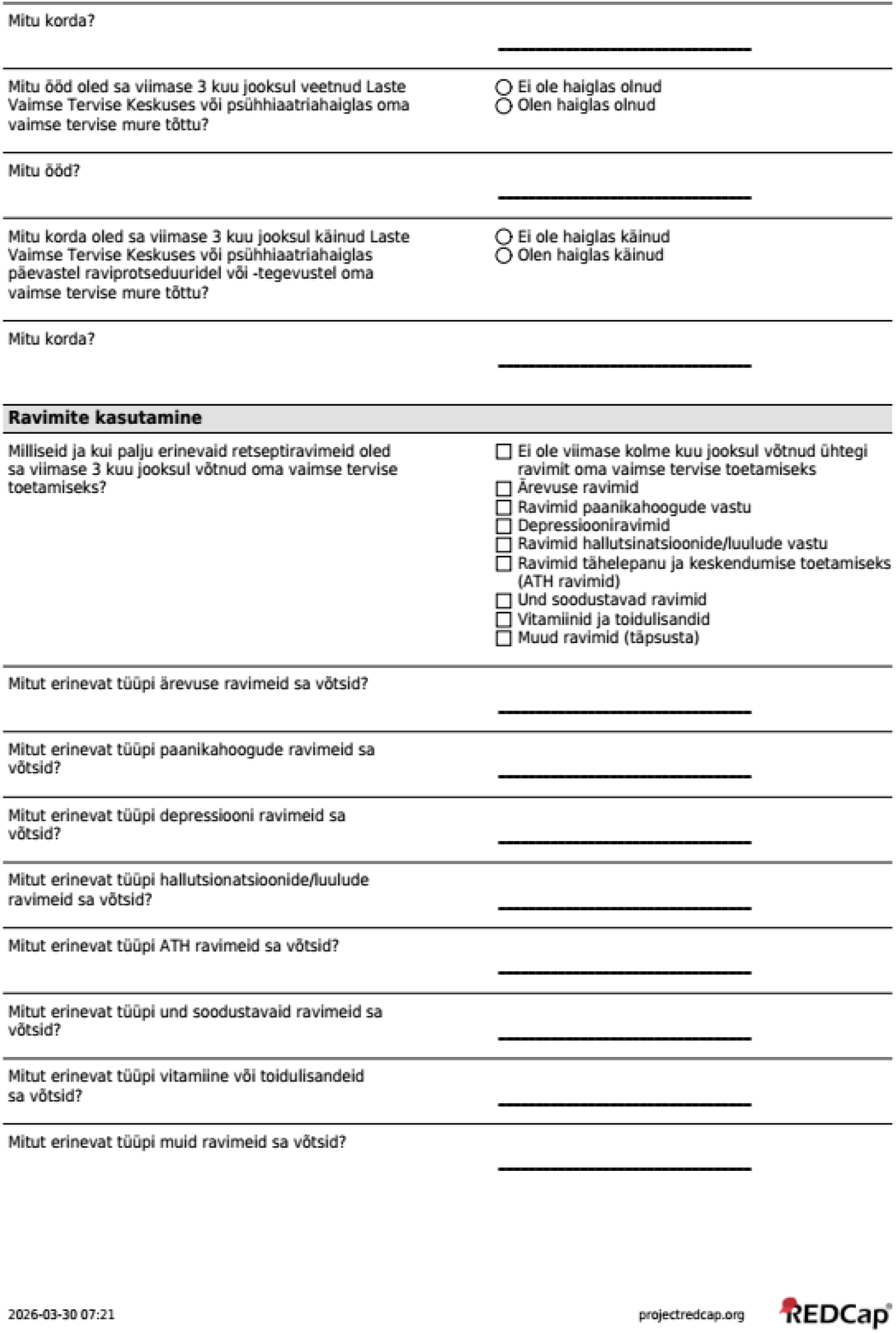
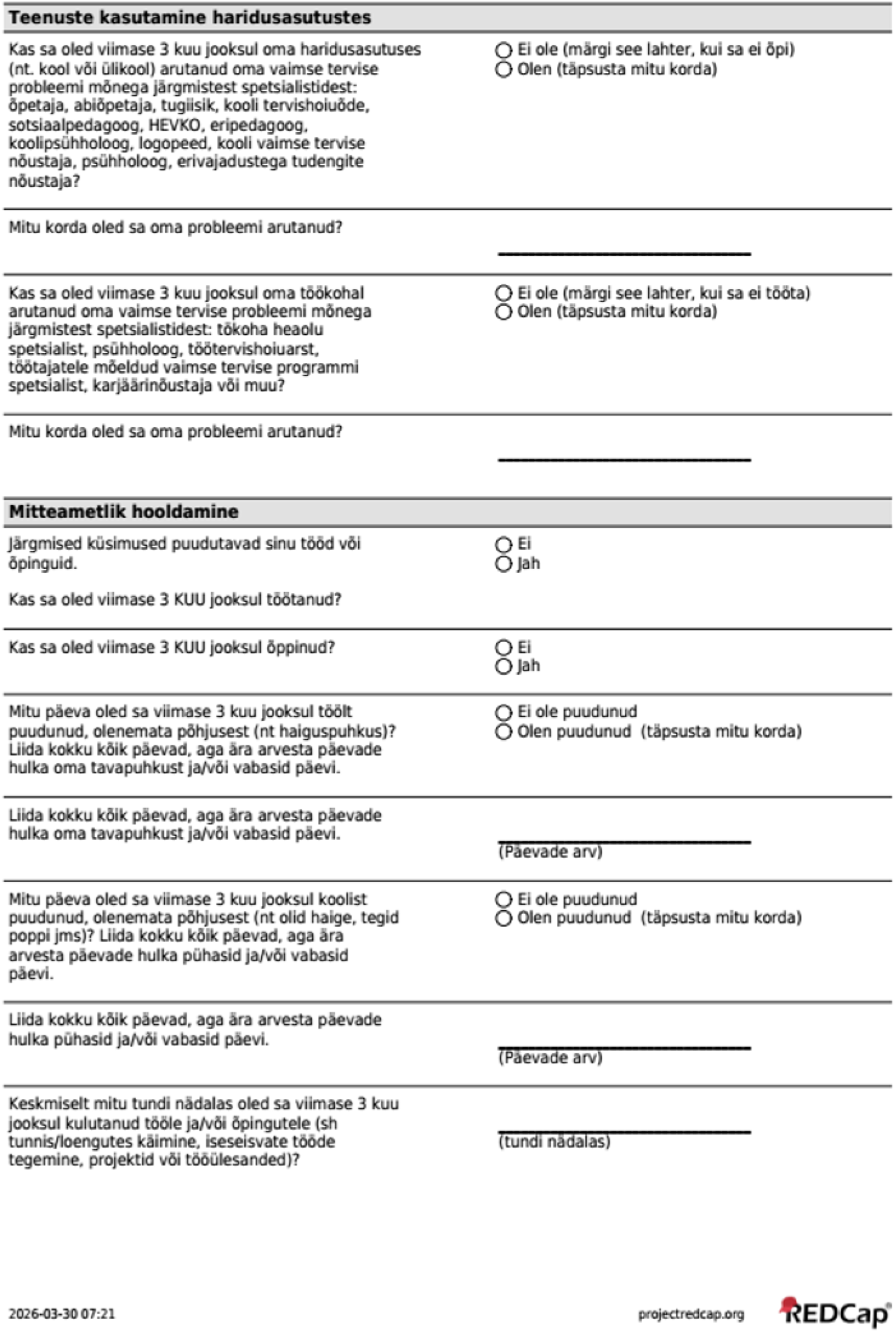
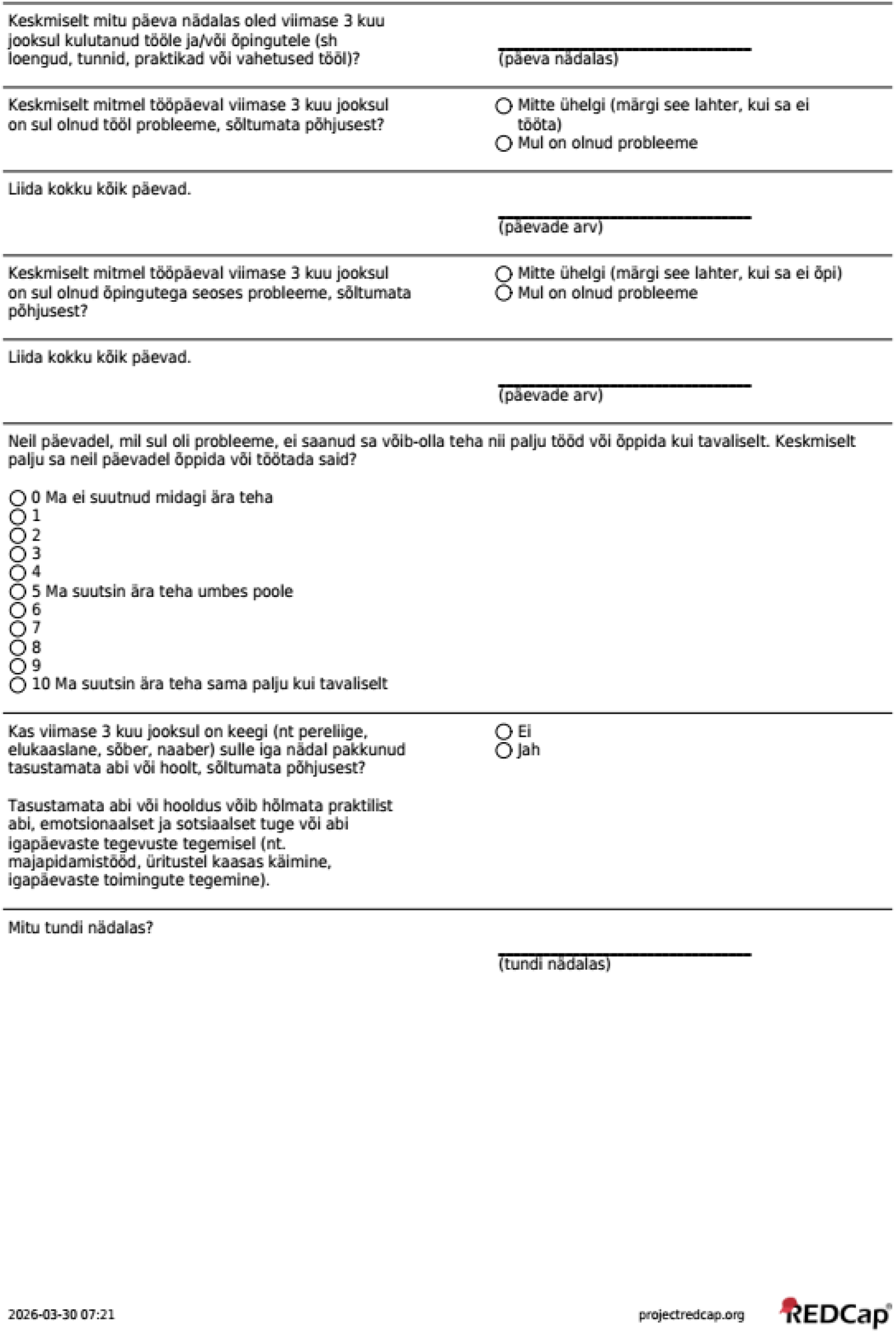

